# Smartphone-based detection of memory decline in prodromal Alzheimer’s disease

**DOI:** 10.1101/2025.06.10.25329344

**Authors:** Sarah E. Polk, Lindsay R. Clark, Kristin Basche, Luca Kleineidam, Wenzel Glanz, Michaela Butryn, Robert Perneczky, Katharina Buerger, Klaus Fliessbach, Christoph Laske, Annika Spottke, Anja Schneider, Jens Wiltfang, Stefan Teipel, Claudia Bartels, Ayda Rostamzadeh, Daniel Janowitz, Boris-Stephan Rauchmann, Ingo Kilimann, Sebastian Sodenkamp, Marie Coenjaerts, Frederic Brosseron, Michael Wagner, Ingo Frommann, Melina Stark, Matthias Schmid, Björn H. Schott, Sterling C. Johnson, Frank Jessen, Emrah Düzel, David Berron

**Affiliations:** German Center for Neurodegenerative Diseases (DZNE), Magdeburg, Germany; Division of Geriatrics, Department of Medicine, University of Wisconsin School of Medicine and Public Health, Madison, Wisconsin, USA; Wisconsin Alzheimer’s Disease Research Center, Madison, Wisconsin, USA; Geriatric Research, Education and Clinical Center, William S. Middleton Memorial Veterans Hospital, Madison, Wisconsin, USA; German Center for Neurodegenerative Diseases (DZNE), Bonn, Germany; Department for Cognitive Disorders and Old Age Psychiatry, University Hospital Bonn, Bonn, Germany; German Center for Neurodegenerative Diseases (DZNE), Munich, Germany; Department of Psychiatry and Psychotherapy, University Hospital, LMU Munich, Munich, Germany; Munich Cluster for Systems Neurology (SyNergy), Munich, Germany; Ageing Epidemiology Research Unit (AGE), School of Public Health, Imperial College London, London, UK; Institute for Stroke and Dementia Research (ISD), University Hospital, LMU Munich, Munich, Germany; Department of Old Age Psychiatry and Cognitive Disorders, University Hospital Bonn and University of Bonn, Bonn, Germany; German Center for Neurodegenerative Diseases (DZNE), Tübingen, Germany; Section for Dementia Research, Hertie Institute for Clinical Brain Research and Department of Psychiatry and Psychotherapy, University of Tübingen, Tübingen, Germany; Department of Neurology, University of Bonn, Bonn, Germany; German Center for Neurodegenerative Diseases (DZNE), Goettingen, Germany; Department of Psychiatry and Psychotherapy, University Medical Center Goettingen, University of Goettingen, Goettingen, Germany; Neurosciences and Signaling Group, Institute of Biomedicine (iBiMED), Department of Medical Sciences, University of Aveiro, Aveiro, Portugal; German Center for Neurodegenerative Diseases (DZNE), Rostock, Germany; Department of Psychosomatic Medicine, Rostock University Medical Center, Rostock, Germany; Department of Psychiatry, University of Cologne, Medical Faculty, Cologne, Germany; Sheffield Institute for Translational Neuroscience (SITraN), University of Sheffield, Sheffield, UK; Department of Neuroradiology, University Hospital LMU, Munich, Germany; Institute for Medical Biometry, Informatics and Epidemiology, University Hospital Bonn, Bonn, Germany; Leibniz Institute for Neurobiology, Magdeburg, Germany; Excellence Cluster on Cellular Stress Responses in Aging-Associated Diseases (CECAD), University of Cologne, Cologne, Germany; Institute of Cognitive Neurology and Dementia Research (IKND), Otto-von-Guericke University, Magdeburg, Germany; Center for Behavioral Brain Sciences, Otto-von-Guericke University Magdeburg, Magdeburg, Germany; Clinical Memory Research Unit, Department of Clinical Sciences, Lund University, Lund, Sweden

## Abstract

The slow progression of Alzheimer’s disease (AD) poses a challenge for the rapid and individual quantification of early disease-driven cognitive decline. Here, we show that frequently administered remote and unsupervised digital cognitive assessments can detect cognitive decline within 30 weeks in early AD. The sample comprised 202 individuals (52–85 years old), who were cognitively unimpaired (CU) or had mild cognitive impairment (MCI). Participants self-administered remote tasks testing object and scene memory precision, associative memory, and familiarity-dependent memory. A short-term decline in the familiarity-dependent task was observed in all patients with an MCI diagnosis, while both the familiarity-dependent task and memory precision for objects were sensitive to decline in amyloid-positive MCI patients specifically. Change in the remote familiarity-dependent task was correlated with multi-year change on annual in-person neuropsychological assessments. In conclusion, frequent remote cognitive testing is a promising tool to feasibly capture and monitor subtle and short-term cognitive decline.

---

Memory decline is one of the earliest observed clinical symptoms related to Alzheimer’s disease (AD) pathology^1^, with patient and caregiver reports of progressive memory loss being core diagnostic components of the clinical syndrome. The speed of memory deterioration is also a leading cause of worry for AD patients^2^. Slowing the rate of memory decline is therefore a crucial goal for disease modifying treatments, and the rate of cognitive change is a key outcome measure in clinical trials, especially in earlier disease stages^3–6^. However, despite being a primary target for interventions, it is still not common practice to quantify the rate of memory decline for use in clinical decision-making. A primary reason for this is the lack of tools that can reliably measure decline within reasonable time periods appropriate for timely clinical management.

Suitable cognitive tools could be optimized to detect early cognitive decline in AD through two approaches. Firstly, clinician-administered neuropsychological tests are typically performed every six to 12 months. Increasing the frequency of measurement would improve the reliability of cognitive tests, thereby improving their sensitivity to change^7^. Secondly, memory function is not monolithically compromised in early AD, but rather those component processes that rely on brain regions in which tau pathology accumulates and which show early signs of neurodegeneration are those that show earliest decline^8^. Thus, a tool that targets those processes which are expected to deteriorate earliest may be more sensitive to cognitive decline in early AD.

The advent of remote digital cognitive testing via individual’s own devices has greatly facilitated the frequent administration of cognitive tests (see refs. ^9,10^ for reviews) in a decentralized manner^11^. In contrast to clinician-administered cognitive tests, it is feasible to administer remote and unsupervised cognitive tests up to multiple times a day in older adults^12–14^. Increasing test frequency theoretically improves the characterization of intra-individual variability, reducing the overall susceptibility to measurement error and improving reliability, which in turn improves the measurement of true cognitive change^7^.

Regarding the specificity of cognitive tests to certain memory processes, many commonly used neuropsychological tests broadly assess memory decline as a general construct. However, given what is known about spatiotemporal progression of AD pathologies, namely the accumulation of beta-amyloid (Aβ) plaques and tau neurofibrillary tangles^15,16^, it is hypothesized that specific episodic memory processes are affected sequentially in early AD, aligning with the progression of pathology^8,17^. Briefly, the progression of neurofibrillary tangles follows a stereotypical spatiotemporal pattern, accumulating first in the (trans-)entorhinal cortex and hippocampus in the medial temporal lobe, and later progressing towards the lateral temporal and parietal cortices^18,19^. The progression of Aβ plaques follows a more diffuse pattern, starting in the neocortex and later affecting other cortical regions, including the entorhinal cortex and hippocampus, subcortical regions, and the brainstem^20–23^.

To implement both of these approaches in the current study, we frequently administered three non-verbal visual memory tasks that recruit brain regions known to be affected by AD pathology. The Object-in-Room Recall (ORR) task was designed to recruit pattern completion, or the neural process by which partial memories are completed from previously experienced episodes, which relies on the hippocampal *cornu ammonis* 3 (CA3)^24^; successful pattern completion supports associative memory. The Mnemonic Discrimination Task for Objects and Scenes (MDT-OS) was designed to tap into pattern separation, or the neural process by which similar stimuli are separated into distinct representations^25–27;^ successful pattern separation promotes greater memory precision. This task has been shown to activate a posterior-medial network for scenes, which includes superior occipital and parietal regions, posterior midline regions, and medial temporal regions including the parahippocampal cortex, as well as an anterior-temporal network for objects, which includes occipital and inferior parietal regions, lateral temporal regions, and the perirhinal cortex^28–30^. Finally, the Complex Scene Recognition (CSR) task was designed to measure recognition memory, reliant on both recollection as well as on familiarity. The CSR recruits a larger network of brain regions, including the medial temporal lobe and frontal and parietal regions^31–33^. Notably, sense of familiarity has been shown to support recognition memory even in patients with bilateral hippocampal injury^34^.

## <u>Aim</u>s and hypotheses of the current study

These three remote and unsupervised tasks capturing associative memory, memory precision, and familiarity-dependent memory were administered to a sample including cognitively unimpaired (CU) individuals and patients with mild cognitive impairment (MCI) weekly or every other week for up to a year. Task performance was analyzed to investigate subtle disease-driven changes in cognition.

We first examined the feasibility of high-frequency remote assessments in the current sample, focusing on adherence and compliance to the assigned schedule.

Next, we were interested whether remote and unsupervised memory assessments are sensitive to subtle cognitive change. We first confirmed that the current sample of MCI patients showed cognitive decline as measured with gold standard in-clinic neuropsychological assessments; we characterized yearly rates of cognitive decline spanning up to eight years and tested for differences in performance on a latent measure similar to the Preclinical Alzheimer’s Cognitive Composite (PACC), which is sensitive to differences in long-term change between cognitively healthy individuals with and without biomarker evidence for AD^35,36^. We then tested whether this cognitive decline was captured by the remotely administered episodic memory tests, hypothesizing that MCI patients would show worse performance at baseline as well as greater decline in performance over time. Additionally, we stratified the MCI group according to Aβ status, hypothesizing that those patients with MCI due to AD (MCI Aβ+) would perform worse and decline more steeply than those with other-cause MCI (MCI Aβ−). During the prodromal stage of AD (MCI due to AD), the medial temporal lobe already shows tau PET uptake, while the neocortex starts to show moderate tau PET uptake^37^. We therefore hypothesized that performance on the ORR and MDT-OS would already be impaired in the current MCI sample, especially in those with amyloid positivity, while performance on the CSR (familiarity-dependent memory) would begin to decline during this stage as the neocortex, including posterior parietal areas, begins to be affected.

Finally, we examined the associations between change in remote task performance and established in-person neuropsychological measures. We also investigated whether in-person tests were sensitive to short-term cognitive changes in a sub-sample of participants with two in-person assessments within one year.

## Methods

### Participants and study design

Volunteers aged 50 years or older were recruited for an unsupervised and remote study (mobile add-on study) through three longitudinal observational cohort studies (DZNE: Longitudinal Cognitive Impairment and Dementia Study [DELCODE]^38^, Wisconsin Registry for Alzheimer’s Prevention [WRAP]^39^, Wisconsin Alzheimer’ Disease Research Center [ADRC]) as well as from the local memory clinic. Details about each of the cohorts can be found in the Supplementary Methods. Participants had to own an app-compatible smartphone or tablet with network access and be able to operate the device without assistance. Those with dementia, sensory or motor impairments, or a neurological illness that would impair their ability to complete the tasks were excluded. The mobile add-on study was approved by the ethics committees of each participating site (the medical faculties at the University of Bonn, Ludwig-Maximilians-University Munich, University of Tübingen, Rostock University, University of Göttingen, and University of Cologne; University of Wisconsin-Madison Institutional Review Board; University Hospital Magdeburg).

### In-clinic assessments

#### Cognitive status

DELCODE and memory clinic participants were categorized as CU without concerns, or were diagnosed with subjective cognitive decline (SCD) or MCI. An SCD diagnosis was given if a patient presented to a memory clinic with memory complaints, but scored above −1.5 SD on the subtests of the Consortium to Establish a Registry for Alzheimer’s Disease (CERAD) neuropsychological test battery based on an age-, sex-, and education-normed performance range^38,40^. MCI diagnoses were made based on the current research criteria by the National Institute on Aging and Alzheimer’s Association (NIA-AA)^41,42^. In the current analyses, those with SCD are considered CU, as they show no clinically confirmed impairment. WRAP and Wisconsin ADRC participants in the mobile add-on study were classified as CU or received an MCI diagnosis based on expert consensus conference review also using NIA-AA criteria^41,42^.

#### Aβ status

Aβ status was determined with available data closest to the start of the mobile add-on study (before or after) for all participants. Briefly, for participants recruited through DELCODE, Aβ status was determined using lumbar cerebrospinal fluid (CSF; Mesoscale Diagnostics LLC, Rockville, USA) Aβ_42_/Aβ_40_ ratio (Aβ+ ≤ 0.08) or, if CSF was unavailable, the individual probability of Aβ positivity in CSF was calculated using age, APOE genotype, and plasma Aβ_42_/Aβ_40_ ratio (Lumipulse panel; Fujirebio Inc., Tokyo, Japan; see ref. ^43^) following ref. ^44^ (Aβ+ > .639). For participants recruited through the memory clinic, Aβ status was determined using CSF Aβ_42_/Aβ_40_ ratio (Lumipulse panel; Fujirebio Inc., Tokyo, Japan; Aβ+ ≤ 0.69). For participants recruited through WRAP and Wisconsin ADRC, Aβ status was determined using CSF Aβ_42_/Aβ_40_ ratio (NeuroToolKit panel; Roche Diagnostics International Ltd, Rotkreuz, Switzerland; Aβ+ ≤ .046) or [^11^C]Pittsburgh compound B (PiB) PET (Aβ+ ≥ 1.19 DVR) if CSF was not available (see ref. ^45^). See Supplementary Methods for details.

#### Neuropsychological testing

Cognitive test batteries were administered annually or biennially by a clinician or other qualified study team member. These included the Mini Mental State Examination (MMSE)^46^ or the Montreal Cognitive Assessment (MoCA)^47^, category and/or phonemic fluency tests, the Alzheimer’s Disease Assessment Scale—Cognitive (ADAS-Cog) Word Recall^48^, the Free and Cued Selected Reminding Test (FCSRT96)^49^, the Rey Auditory Verbal Learning Test^50^, the Symbol Digit Modalities Test (SDMT)^51^, the Trail Making Test (TMT)^52^, the Wechsler Adult Intelligence Scale—Revised (WAIS-R) Digit Symbol test (DST)^53^, and the Logical Memory subscale from the Wechsler Memory Scale—Revised and Fourth Versions (WMS-R and WMS-IV)^54,55^ or the Craft Story 21^56^. See Supplementary Methods for details.

To harmonize neuropsychological test data across cohorts, a confirmatory factor analysis was performed using structural equation modeling with the *lavaan* package^57^ to determine the loadings of each individual test onto a latent PACC (lPACC) factor at the chronologically closest in-person study visit to the start of the mobile add-on study. This model showed acceptable fit indices, CFI = .912, RMSEA = .069. These loadings were then used in a second step to extract lPACC scores at every time point for each individual (see Supplementary Methods for more details) for use in subsequent analyses.

#### Mobile add-on study

Participants completed remote and unsupervised cognitive testing using the neotivTrials app (neotiv GmbH) on their own smart devices for at least 30 weeks. All participants were assisted in downloading the app onto their smartphone or tablet, either in person or via email and phone. Each participant received a pseudonymized ID to use in the app, so that no personal or clinical information was associated with their app data. All data were transferred to the appropriate research centers in accordance with the General Data Protection Regulation. Remote data were then merged with clinical data before being made available to the principal investigators involved in the current analyses.

Participants recruited through DELCODE and 38 participants recruited through WRAP/Wisconsin ADRC were instructed to complete one task every two weeks, while those recruited through the memory clinic completed one task every week. The remaining 53 participants recruited through WRAP and Wisconsin ARDC completed all three tasks in a multi-day burst every two months. As all participants from WRAP and Wisconsin ADRC were assigned the same mobile add-on study schedule for each task respectively, they are subsequently referred to as the Wisconsin cohort.

Participants completed three tasks in the app. The MDT-OS (memory precision) is a lure discrimination task with 32 object and 32 scene items with one- and two-back phases. The corrected hit rate, that is, the hit rate (percentage of correctly identified repeat items) minus the false alarm rate (percentage of incorrectly identified lure items), was calculated across phases for objects (MDT-O) and scenes (MDT-S) separately, and the average corrected hit rate between phases was used in subsequent analyses. The ORR (associative memory) is a 25-item room-object-location association task with immediate and delayed recall phases. The average score between phases was calculated and used in subsequent analyses, and time elapsed between the phases was included in the models as a nuisance variable. Finally, the CSR (familiarity-dependent memory) is a delayed recognition task with 60 photographic images depicting scenes. The corrected hit rate was calculated and used in subsequent analyses, and time elapsed between the learning and recognition phases was taken into account. After each task, participants were asked if they had been distracted during the task (yes/no), and how well they concentrated during the task (0 to 4, with 4 being the highest). See Fig. 1 for a visual representation of the study design and see Supplementary Methods for more details regarding the tasks.

**Fig. 1.**
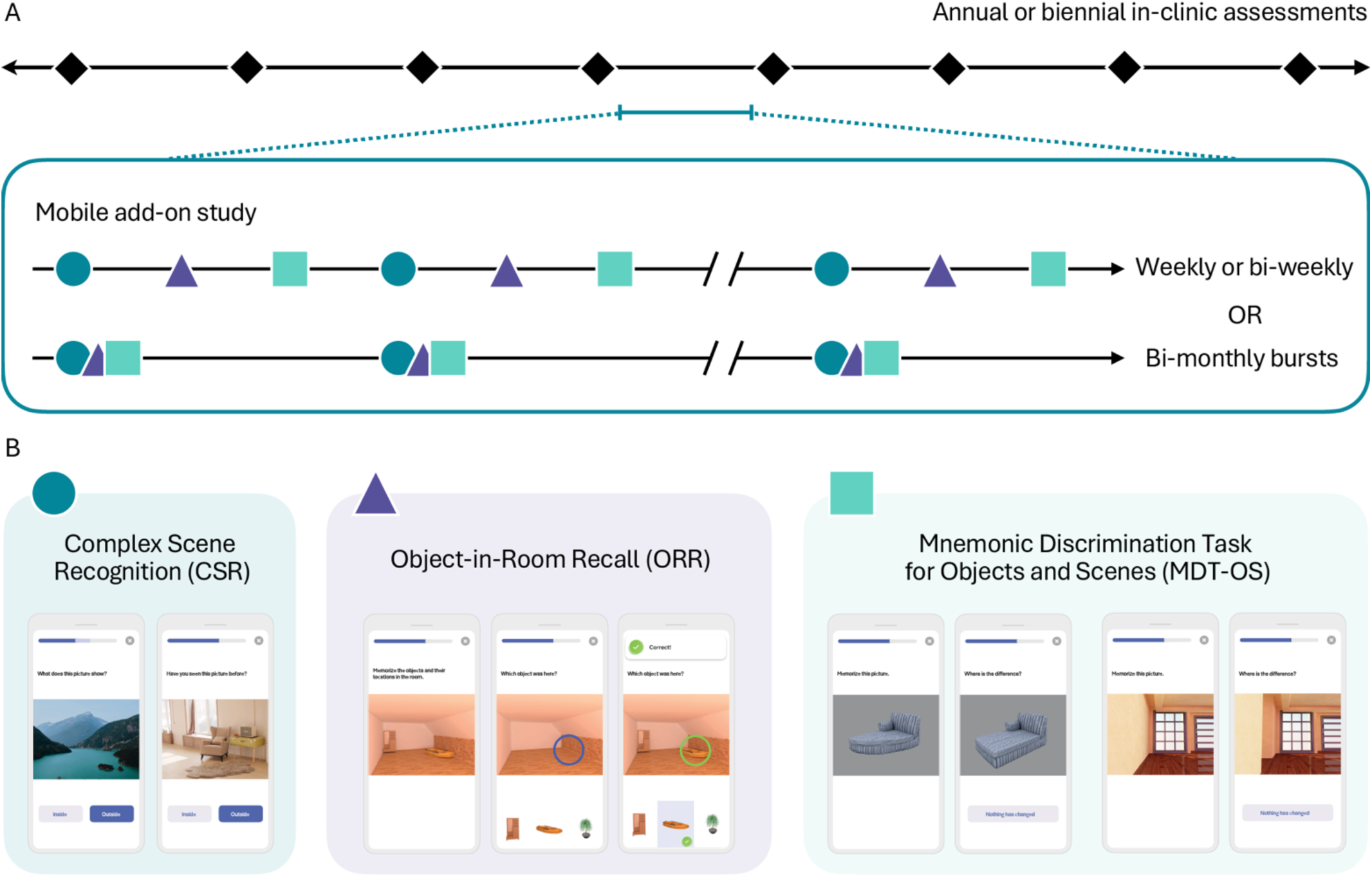
Study design. (A) Participants were recruited from ongoing parent studies with annual or biennial in-clinic assessments (diamonds) to participate in the mobile-add on study in one of two possible designs: a continuous design with weekly or bi-weekly remote assessments or a burst design with four-day bursts every second month. (B) The tasks included a task for familiarity-dependent memory (Complex Scene Recognition or CSR; circles), a task for associative memory (Object-in-Room Recall or ORR; triangles), and a task for memory precision of objects and scenes (Mnemonic Discrimination Task for Objects and Scenes or MDT-OS; squares).

#### Task data processing

To ensure data quality, a number of filtering steps were applied to the task data before analyses. These steps included removing data points that were unusable due to technical problems, prolonged time to retrieval (i.e., the time elapsed between phases for ORR and CSR), and excessive timeouts (i.e., unfinished trials). See Supplementary Methods for details on how many data points were excluded during the filtering process. After filtering, data were standardized such that the CU group had a mean of 0 and standard deviation of 1 at baseline.

#### Statistical analyses

All data wrangling, statistical analysis, and visualization was done using R in RStudio^58,59^ with the *tidyverse* package^60^. An alpha of .05 was used to test for statistical significance. In the current analyses, we were interested in cognitive changes in MCI, particularly in MCI due to AD. Therefore, we included only those CU individuals with normal Aβ levels (CU Aβ−) in the healthy control group, comparing them to either all MCI patients, or stratifying MCI patients by Aβ burden (MCI Aβ− and MCI Aβ+). Those MCI patients for whom the Aβ status was unknown were excluded from the second set of analyses; the samples were otherwise identical.

#### Adherence and compliance

To test the feasibility of high-frequency remote assessments in the current sample, adherence and compliance to the mobile add-on study were calculated for each individual and compared across cognitive status groups and study cohorts. Here, we define adherence as the number of completed task sessions divided by the maximum number of possible sessions in a given study protocol (percentage of tasks completed). Compliance was calculated as the number of task sessions completed divided by how many tasks an individual could have completed given how long they participated in the mobile add-on study (e.g., if a participant from the memory clinic was in the study for 10 weeks, they should have completed 10 tasks). Therefore, it is possible for a participant to have imperfect adherence but perfect compliance or *vice versa* (see Supplementary Methods for examples). Two-way ANCOVAs were used to investigate differences in adherence and compliance across cognitive status (CU, MCI) and study cohort (DELCODE, memory clinic, Wisconsin), taking age, sex, and years of education into account. *Post hoc* pairwise comparisons were done using the *emmeans* package^61^.

#### Confirmatory analyses of decline in in-person neuropsychological assessments

Before addressing our second aim regarding performance on the remote and unsupervised digital cognitive tests, we first wanted to establish the yearly rate of cognitive decline within the current sample using gold standard measures, in this case the harmonized lPACC. To this end, linear mixed models (LMMs) were used to analyze the lPACC scores, controlling for age at the start of the mobile add-on study, sex, years of education, and study cohort (DELCODE, memory clinic, Wisconsin). LMMs including group (CU or MCI) and all covariates as fixed effects, time spent in the study (centered at each participant’s first remote assessment) as both a fixed and random effect, individual intercept as a random effect, and a group-by-time interaction term were built. Participants with only one time point were excluded from the analyses. The model was estimated using the maximum likelihood estimator and the BOBYQA optimizer^62^. Main effect of group (i.e., group differences at intercept) and group-by-time interaction (i.e., group differences in slope) are reported.

#### Differences in remote task performance between diagnostic groups and the effects of Aβ positivity

To address our second aim related to the remote tasks, LMMs were also used to investigate the difference in change in task performance over 30 weeks between individuals with CU and MCI, controlling for age at the start of the mobile add-on study, sex, years of education, and study cohort (DELCODE, memory clinic, Wisconsin), as well as time to retrieval for ORR and CSR, distraction (yes/no), and concentration (0–4), following the same steps as described above. Additionally, to account for differences in study design across the cohorts and potentially differential effects of exposure to the task, we also controlled for task session (1–10) as a factor. Participants with only one time point were excluded from the analyses. Separate models were used for each task (MDT-O, MDT-S, ORR, CSR). Within-group intercepts and slopes were estimated and compared pairwise using the *emmeans* package^61^.

Similar LMMs as described above were used to investigate the difference in change in task performance over 30 weeks, stratifying the MCI group into MCI Aβ− and MCI Aβ+ groups, such that each sub-group was compared to the CU group. The same covariates were included in the models, and the same analysis steps were carried out, with separate models for each task.

#### Comparisons of remote and in-person cognitive tests

To understand the relationship between change in performance on the remotely assessed cognitive tests and those conducted in person, slope-slope correlations were calculated. We first tested for variance in change in all variables (lPACC and remote tasks) by comparing two models with different random effect structures: 1) including both random intercepts and slopes and 2) including only random intercepts. If the random-slopes model showed significantly improved fit over the random-intercept model, random slopes (i.e., change estimated for each individual) were extracted from LMMs including time as a fixed and random effect and the aforementioned covariates (no group term). Random slopes on in-person neuropsychological assessments were correlated with random slopes on remote task performance using the Pearson method across groups as well as within each diagnostic group.

Finally, to investigate whether in-person neuropsychological assessments are also sensitive to changes over a shorter time frame (i.e., one year), the lPACC score from the in-person appointments closest to the start of the mobile add-on study as well as the subsequent one (if the two assessments fell within one year of each other) were included in an LMM accounting for age, sex, education, and study cohort, modeling random intercepts.

## Results

### Participants

We included 202 eligible participants from the remote mobile add-on study in the current analysis, comprising 152 CU participants and 50 MCI patients. Sample characteristics and summaries of key measures at baseline can be found in Table 1.

**Table 1.** Participant demographics, information about remote testing, and baseline scores on key measures. CU = cognitively unimpaired, MCI = mild cognitive impairment, MMSE = Mini-Mental State Examination, CDR = Clinical Dementia Rating, lPACC = latent Preclinical Alzheimer’s Cognitive Composite, MDT-O/-S = Mnemonic Discrimination Task for Objects/Scenes (corrected hit rate), ORR = Object-in-Room Recall (score out of 25), CSR = Complex Recognition Task (corrected hit rate). *N*s and percentages of all participants in the (sub-) sample are reported for Participants, Aβ status, Study cohort, and Sex. Means and standard deviations are reported for all other variables. Age was calculated at each participant’s first remote task session. MMSE, CDR Sum of Boxes, and lPACC from the in-person assessment chronologically closest to the start of the mobile add-on study are reported. Results from an ANCOVA controlling for age, sex, and years of education are reported for the MMSE, CDR Sum of Boxes, lPACC, and remote task performance; time to retrieval was also included for ORR and CSR. Note that CDR Sum of Boxes was not available for Memory clinic participants. * *p* < .050 indicating a significant difference across groups.

|  | All | CU | MCI | Group difference |
| --- | --- | --- | --- | --- |
| Participants | 202 | 152 (75%) | 50 (25%) |  |
| Aβ status | 5 missing | 0 missing | 5 missing | |
| Aβ- | 173 (86%) | 152 (100%) | 21 (42%) | |
| Aβ+ | 24 (12%) | — | 24 (48%) | |
| Study cohort |  |  |  |  |
| DELCODE | 77 (38%) | 60 (39%) | 17 (34%) |  |
| Memory clinic | 34 (17%) | 4 (3%) | 30 (60%) | $\chi^2_{2, 202} = 96.35^*$ |
| Wisconsin | 91 (45%) | 88 (58%) | 3 (6%) |  |
| Age (years) | 69.9 ± 6.8 | 69.2 ± 6.6 | 72.1 ± 7.1 | $F_{1, 200} = 7.10^*$ |
| Sex (female) | 118 (58%) | 98 (64%) | 20 (40%) | $\chi^2_{1, 202} = 8.30^*$ |
| Education (years) | 15.4 ± 2.7 | 15.9 ± 2.5 | 13.8 ± 2.6 | $F_{1, 200} = 25.98^*$ |
| MMSE | 28.5 ± 2.6 | 29.5 ± 0.7 | 25.8 ± 3.8 | $F_{1, 183} = 122.46^*$ |
| CDR Sum of Boxes | 0.2 ± 0.5 | 0.1 ± 0.3 | 0.7 ± 0.9 | $F_{1, 147} = 29.72^*$ |
| Number of remote sessions completed | 22.6 ± 10.3 | 22.5 ± 8.8 | 22.8 ± 14.0 | $F_{1, 197} = 0.03$ |
| Time spent in mobile add-on study (weeks) | 39.6 ± 20.6 | 42.7 ± 19.2 | 30.1 ± 21.9 | $F_{1, 197} = 15.04^*$ |
| Baseline performance |  |  |  |  |
| IPACC | -0.64 ± 1.32 | -0.06 ± 0.73 | -2.36 ± 1.16 | $F_{1, 194} = 314.50^*$ |
| Memory precision for objects (MDT-O) | 0.51 ± 0.20 | 0.54 ± 0.18 | 0.39 ± 0.21 | $F_{1, 171} = 22.27^*$ |
| Memory precision for scenes (MDT-S) | 0.42 ± 0.21 | 0.48 ± 0.18 | 0.24 ± 0.19 | $F_{1, 171} = 58.29^*$ |
| Associative memory (ORR) | 19.7 ± 2.7 | 20.4 ± 2.2 | 16.1 ± 2.3 | $F_{1, 152} = 83.07^*$ |
| Familiarity-dependent memory (CSR) | 0.53 ± 0.20 | 0.58 ± 0.18 | 0.35 ± 0.18 | $F_{1, 170} = 57.62^*$ |

### Adherence and compliance to the remote and unsupervised tests is acceptable

To ensure that frequent administration of remote and unsupervised digital cognitive assessments was feasible in the current sample, we examined adherence and compliance to the study schedule (see Fig. 2). Overall, there was an average adherence rate of 78% (SD = 33%) and compliance rate of 90% (SD = 17%). A two-way ANCOVA accounting for age, sex, and years of education revealed a significant main effect of cognitive status on adherence, *F*_1, 190_ = 15.05, *p* < .001, as well as on compliance, *F*_1, 190_ = 20.67, *p* < .001. A main effect of cohort was also observed, adherence: *F*_2, 190_ = 4.31, *p* = .015, compliance: *F*_2, 190_ = 5.96, *p* = .003. Grouped pairwise comparisons showed no significant differences in adherence. Within DELCODE participants, the CU group showed better compliance than the MCI group, and CU participants from the Wisconsin cohort showed better compliance than CU participants from DELCODE.

**Fig. 2.**
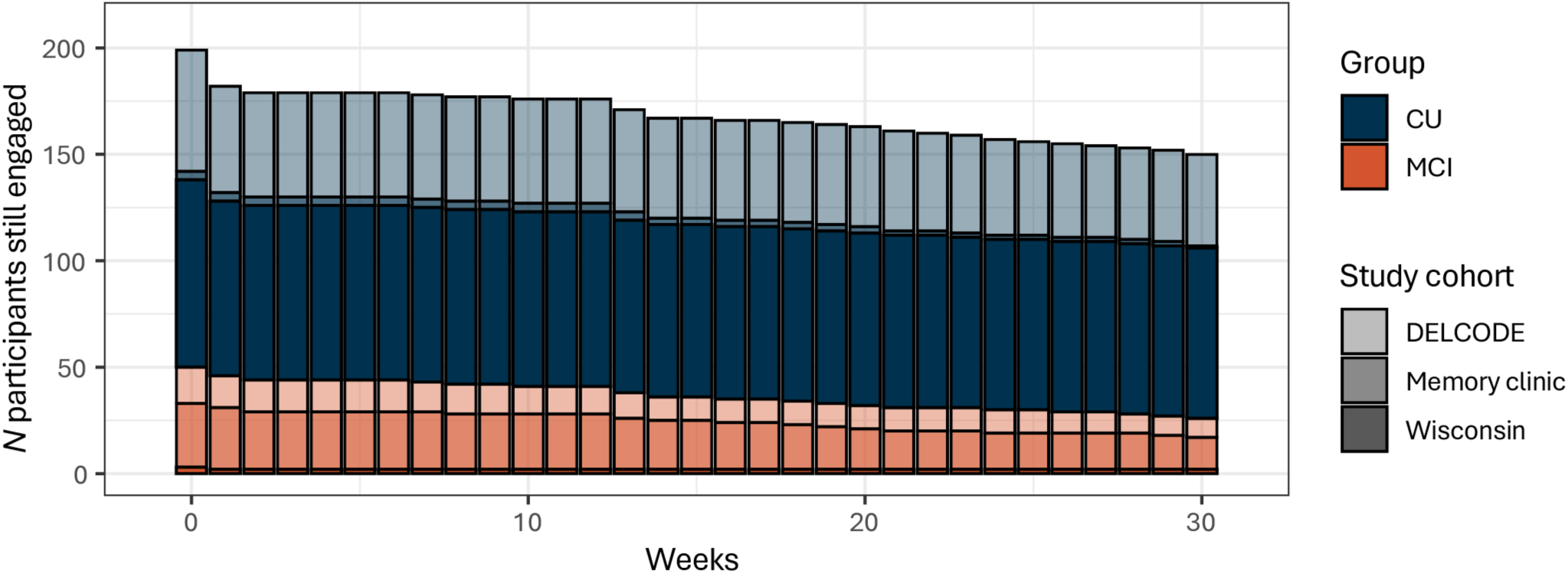
Adherence to the mobile add-on study indicated good feasibility. Out of the 202 participants from the mobile add-on study included in these analyses, 199 had valid task data at baseline. By week 30, 150 participants (74%) were still actively engaged in the mobile add-on study. CU = cognitively unimpaired; MCI = mild cognitive impairment.

### Characterization of long-term cognitive decline with in-person neuropsychological tests

Before addressing our aims regarding performance on the remotely administered cognitive tasks, we first characterized annual cognitive decline measured with gold standard metrics, in this case the cross-cohort harmonized lPACC, using LMMs. When comparing CU participants and all MCI patients, the MCI group performed worse at baseline, *b* = −1.455, 95% CI [−2.150, −1.615], and showed a significantly more negative slope, *b* = −0.145, 95% CI [−0.189, −0.102]. When stratifying the MCI group according to Aβ status, significant baseline differences were observed for the MCI Aβ+ group, *b* = −0.246, 95% CI [−0.302, −0.193], but not the MCI Aβ− group, *b* = −0.048, 95% CI [−0.103, 0.005] (see Fig. 3). See Supplementary Tables 3 and 4 for full results.

**Fig. 3.**
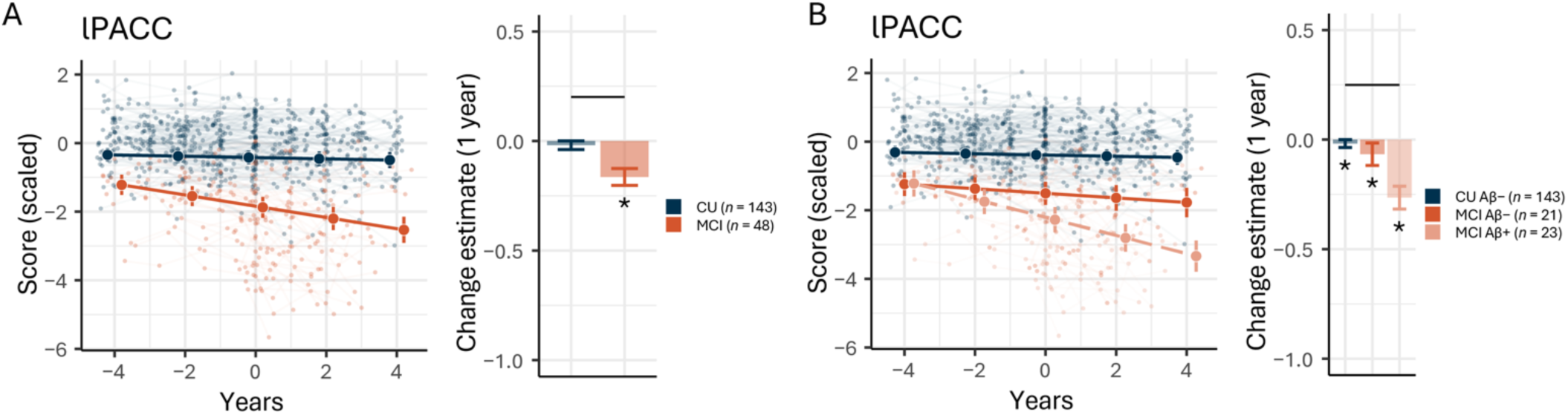
Multi-year change in lPACC. (A) Patients with a mild cognitive impairment (MCI) diagnosis (orange) showed signficantly greater decline in latent Preclinical Alzheimer’s Cognitive Composite (lPACC) scores than cognitively unimpaired (CU) individuals (dark blue) over eight years, centered at the start of the mobile add-on study. (B) When stratifying according to amyloid-β (Aβ) status, both Aβ− (orange) and Aβ+ (peach-colored) patients with MCI showed greater decline than the CU group. Solid black lines indicate differences in change between groups at *p* < .050; asterisks indicate a significant within-group estimate against zero at *p* < .050. Error bars represent 95% confidence intervals.

### Remote and unsupervised assessments capture short-term declines in episodic memory

To capture group differences in performance on the remote tasks, LMMs were run comparing the CU and whole MCI groups, then again stratifying the MCI group by Aβ status. For the sake of visual clarity, all statistics of interest are reported in Table 2. The first comparison revealed baseline differences on all tasks, with all MCI patients performing worse than the CU group at baseline. When stratifying the MCI group according to Aβ status, this pattern remained consistent, such that both MCI sub-groups performed worse than the CU group, with no differences seen between the MCI Aβ− and MCI Aβ+ groups.

**Table 2.** Model estimates of the main effect of group (difference at intercept) on task performance and group-by-time interaction effect (difference in slope), and within-group estimates. Unstandardized estimates and 95% confidence intervals are reported. MDT-O/-S = Mnemonic Discrimination Task for Objects/Scenes, ORR = Object-in-Room Recall, CSR = Complex Scene Recognition, CU = cognitively unimpaired, MCI = mild cognitive impairment. Estimates in bold are significant at *p* < .050.

|  |  | CU vs. all MCI |  |  | CU vs. MCI Aβ- and MCI Aβ+ |  |  |  |  |
| --- | --- | --- | --- | --- | --- | --- | --- | --- | --- |
|  |  | Group differences | Within-group estimates |  | Group differences |  | Within-group estimates |  |  |
| Parameter |  | CU vs. MCI | CU | MCI | CU vs. MCI Aβ- | CU vs. MCI Aβ+ | CU | MCI Aβ- | MCI Aβ+ |
| Memory precision for objects (MDT-O) | Main effect of group | **-0.587**<br>[**-1.056, -0.118**] | 0.487<br>[-0.066, 1.040] | -0.100<br>[-0.826, 0.626] | **-0.667**<br>[**-1.205, -0.127**] | **-0.608**<br>[**-1.162, -0.057**] | **0.675**<br>[**0.125, 1.226**] | 0.009<br>[-0.747, 0.764] | 0.067<br>[-0.720, 0.854] |
|  | Group-by-time interaction | -0.990<br>[-2.069, 0.086] | -0.964<br>[-2.014, 0.086] | **-1.954**<br>[**-3.575, -0.333**] | -0.088<br>[-1.36, 1.180] | **-1.287**<br>[**-2.499, -0.087**] | **-1.115**<br>[**-2.174, -0.056**] | -1.203<br>[-2.971, 0.565] | **-2.401**<br>[**-4.094, -0.709**] |
| Memory precision for scenes (MDT-S) | Main effect of group | **-0.903**<br>[**-1.385, -0.424**] | 0.000<br>[-0.573, 0.573] | **-0.903**<br>[**-1.641, -0.166**] | **-0.999**<br>[**-1.568, -0.432**] | **-0.887**<br>[**-1.469, -0.306**] | 0.104<br>[-0.488, 0.696] | **-0.895**<br>[**-1.698, -0.093**] | -0.783<br>[-1.618, 0.052] |
|  | Group-by-time interaction | 0.823<br>[-0.244, 1.889] | -0.398<br>[-1.506, 0.710] | 0.425<br>[-1.221, 2.071] | 1.235<br>[-0.126, 2.595] | 0.635<br>[-0.656, 1.924] | -0.409<br>[-1.555, 0.737] | 0.826<br>[-1.058, 2.709] | 0.225<br>[-1.577, 2.028] |
| Associative memory (ORR) | Main effect of group | **-1.400**<br>[**-1.898, -0.903**] | 0.046<br>[-0.495, 0.588] | **-1.354**<br>[**-2.076, -0.632**] | **-1.256**<br>[**-1.821, -0.689**] | **-1.847**<br>[**-2.491, -1.206**] | 0.068<br>[-0.482, 0.618] | **-1.188**<br>[**-1.967, -0.408**] | **-1.779**<br>[**-2.619, -0.938**] |
|  | Group-by-time interaction | 0.583<br>[-0.448, 1.616] | -0.191<br>[-1.206, 0.824] | 0.392<br>[-1.263, 2.047] | 0.872<br>[-0.393, 2.133] | 0.750<br>[-0.480, 1.993] | -0.261<br>[-1.298, 0.777] | 0.611<br>[-1.193, 2.415] | 0.489<br>[-1.325, 2.303] |
| Familiarity-dependent memory (CSR) | Main effect of group | **-0.688**<br>[**-1.109, -0.268**] | 0.004<br>[-0.480, 0.488] | **-0.684**<br>[**-1.303, -0.066**] | **-0.823**<br>[**-1.292, -0.355**] | **-0.669**<br>[**-1.183, -0.154**] | -0.115<br>[-0.621, 0.391] | **-0.938**<br>[**-1.614, -0.263**] | **-0.784**<br>[**-1.491, -0.077**] |
|  | Group-by-time interaction | **-1.068**<br>[**-1.963, -0.162**] | -0.121<br>[-1.007, 0.765] | -1.189<br>[-2.591, 0.212] | -0.636<br>[-1.762, 0.511] | **-1.352**<br>[**-2.514, -0.179**] | 0.065<br>[-0.861, 0.991] | -0.571<br>[-2.124, 0.982] | -1.287<br>[-2.924, 0.350] |

Regarding differences in change, the CU and MCI groups showed a difference in familiarity-dependent memory (CSR) performance slope, with the MCI group showing a significantly steeper decline compared to the CU group. The MCI group also showed a significantly negative change in memory precision for objects (MDT-O), but the difference in change between the groups was only marginal, *p* = .071. When stratifying the MCI group into MCI Aβ− and MCI Aβ+ groups, differences in change between the CU and the MCI Aβ+ groups were observed in memory precision for objects and familiarity-dependent memory. No differences in change were seen between the CU and MCI Aβ− groups. *Post hoc* pairwise comparisons did not reveal any significant differences in change between the MCI Aβ− and MCI Aβ+ groups. See Table 2 for group difference estimates as well as within-group estimates and Fig. 4 for a visual representation of the results. See Supplementary Tables 5–12 for full results.

**Fig. 4.**
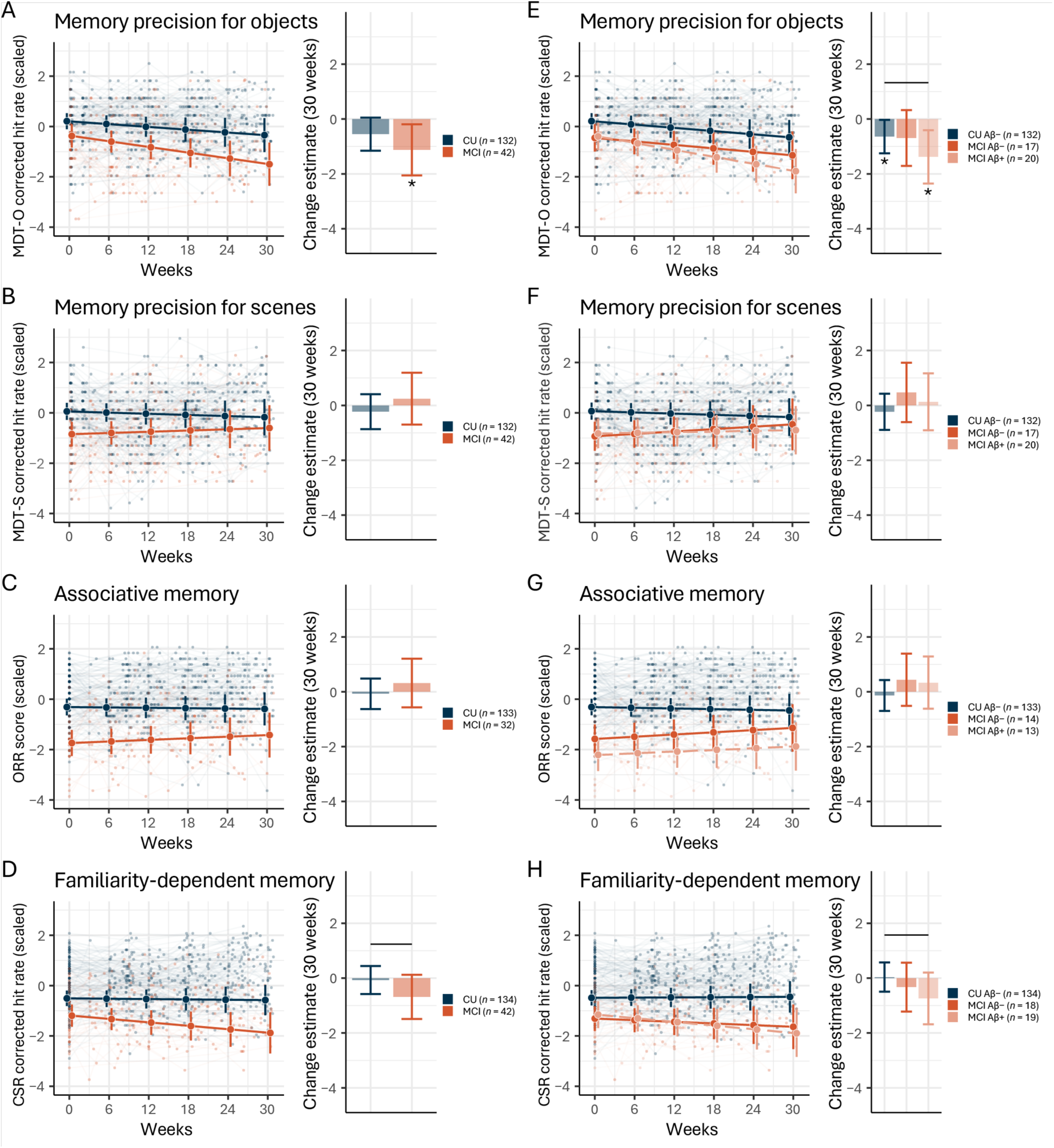
Estimated longitudinal performance trajectories according to cognitive status and stratified by Aβ status. Patients with mild cognitive impairment (MCI) did not differ from cognitively unimpaired (CU) individuals in change in (A) memory precision for objects (MDT-O), (B) memory precision for scenes (MDT-S), or (C) associative memory (ORR), but showed significantly greater decline in (D) familiarity-dependent memory (CSR), indicated by the solid black line. When stratifying the MCI group by Aβ status, the MCI Aβ+ showed greater decline in (E) memory precision for objects (MDT-O), not (F) memory precision for scenes (MDT-S) or (G) associative memory (ORR), and also on (H) familiarity-dependent memory (CSR). Error bars represent 95% confidence intervals. * within-group difference significantly different from 0, *p* < .050.

### Short-term change on remote assessments reflects long-term change on in-person neuropsychological tests

We calculated slope-slope correlations between short-term change tested remotely and multi-year in-person neuropsychological assessments. The associative memory (ORR) and memory precision for scenes (MDT-S) tasks did not exhibit sudicient variance in change to reliably measure covariance, therefore correlations were only investigated for memory precision for objects (MDT-O) and familiarity-dependent memory (CSR). Longitudinal correlations were not significant for memory precision for objects, *r* = −.04, *p* = .586, and were significantly positive for familiarity-dependent memory, *r* = .24, *p* = .002. The CU group showed a significant change-change correlation between the lPACC and familiarity-dependent memory, *r* = .27, *p* = .003, while the MCI group did not, *r* = –.01, *p* = .949. Group-wise correlations between change in lPACC and change in memory precision for objects were not significant, CU: *r* = −.16, *p* = .074, MCI: *r* = −.03, *p* = .835 (see Supplementary Fig. 6).

### In-person neuropsychological tests are not sensitive to short-term changes

To test whether the lPACC could capture short-term cognitive change, two lPACC scores within a year of each other were included in an LMM, *N* = 46 (*n*_MCI_ = 25). On average, the measurements were 39.4 weeks apart (range = 8.0 to 52.1 weeks). When comparing the CU and MCI groups, we observed a difference at baseline, *b* = −1.560, 95% CI [−2.210, −0.905], with the MCI group performing worse than the CU group. The groups did not show a difference in change, *b* = −0.018, 95% CI [−0.467, 0.443]. When stratifying the MCI group by Aβ status (*N* = 45; *n*_MCI Aβ_− = 10, *n*_MCI Aβ+_ = 14), group effects were observed at baseline, with the CU group performing better than both the MCI Aβ− group, *b* = −1.293, 95% CI [−1.956, −0.631], and the MCI Aβ+ group, *b* = −2.172, 95% [−2.887, −1.451]. Change in the MCI Aβ− group was not significantly different from change in the CU group, *b* = 0.261, 95% CI [−0.266, 0.789], nor was change in the MCI Aβ+ group, *b* = −0.266, 95% CI [−0.807, 0.304]. A pairwise comparison showed that change in the MCI Aβ− and MCI Aβ+ groups did not differ significantly. See Supplementary Tables 13 and 14 for full results.

Notably, when running the LMMs for the remote tasks in this restricted sample, we observed results consistent with those found in the whole sample. Namely, a significant difference in change between the CU and MCI groups for memory precision for objects was seen, *b* = −1.674, 95% CI [−3.162, −0.129], as well as a marginal difference in change for familiarity-dependent memory, *b* = −1.265, 95% CI [−2.742, 0.234] (*p* = .095). Stratifying the MCI group by Aβ status also yielded consistent findings, with the MCI Aβ+ group exhibiting a significant difference in change in memory precision for objects when compared to the CU group, *b* = −2.621, 95% CI [−4.398, −0.865], as well as a marginal difference in change for familiarity-dependent memory, *b* = − 1.543, 95% CI [−3.475, 0.378] (*p* = .115).

## Discussion

In the current analyses, we investigated cognitive trajectories in older adults over 30 weeks using frequent remote unsupervised cognitive assessments. First, we established the feasibility of deploying remote and unsupervised cognitive assessments for up to one year. Then, differences between a CU and an MCI group in change on three tests of episodic memory processes were tested and the effect of Aβ status in the MCI group was explored. Finally, we examined the change-change correlations between remotely measured cognitive performance and cognition measured with established in-person neuropsychological assessments (lPACC), and also tested the sensitivity of in-clinic measurements to short-term cognitive change.

### Frequent remote and unsupervised longitudinal cognitive assessments are feasible

Regarding feasibility, we found that, on average, participants completed 78% of all task sessions, indicating that participants completed most of the assigned remote tasks, and average compliance was 90%, showing that the majority of participants, even those who eventually dropped out of the study, completed the tasks in a timely manner. This is comparable to a previous year-long remote study, which had an adherence rate of 86% in a study design with monthly remote tasks^63^. The MCI group showed worse compliance to the study schedule than the CU group; it is plausible that MCI patients may have greater difficulties remembering to complete tasks at a specific time. Future research may take extra care that MCI patients are sent salient reminders to complete tasks on time. Additionally, those CU participants from the Wisconsin cohort showed better compliance than DELCODE participants. This may be because participants from the Wisconsin cohort were contacted if they failed to engage with the study app for a prolonged period of time, while contacting DELCODE participants was not possible. In general, the high adherence and compliance rates indicate that high-frequency remote and unsupervised assessment over up to a year is feasible in older adults with varying degrees of cognitive impairment, though future studies may consider adopting strategies to improve feasibility even further.

### Remote tasks are sensitive to short-term memory decline in all-cause MCI and MCI due to AD

Regarding long-term cognitive change, we found that the MCI patients in the current subsample showed decline on established in-person neuropsychological tests over up to eight years, as expected. In terms of short-term change measured remotely, we found that, while associative memory and memory precision did not show any differences in change between the CU and all-cause MCI groups, performance on the CSR, that is, familiarity-dependent memory, declined in MCI patients compared to CU participants, supporting our hypotheses. Additionally, memory precision for objects and familiarity-dependent memory both showed differences in change between the CU group and MCI Aβ+ group. This suggests that these tasks, when frequently administered, may be suitable tools to monitor cognitive change in MCI, particularly in MCI patients with an elevated amyloid burden, over short periods of time (30 weeks).

Within a moderately sized sub-sample, we found no evidence that in-person tasks are sensitive to short-term decline, even in MCI patients with AD pathology. Namely, when limiting the analyses to only two measurements per person taken within a year of each other, the lPACC score was not sensitive to differences in change between the CU group and the MCI group, even when stratifying according to Aβ status. This further underlines the need for tools that can capture short-term cognitive changes, either by frequently assessing cognition to improve measurement reliability, administering tasks that are specific to AD-related changes in cognition, or a combination of both approaches. Notably, when restricting the analysis of the remote task performance to the same sub-sample, we were still able to detect differences in change in memory precision for objects and familiarity-dependent memory between the CU and MCI groups. Altogether, these findings suggest that frequently administered remote and unsupervised cognitive tasks are an appropriate tool to measure subtle cognitive changes that are otherwise undetectable with in-clinic neuropsychological measures.

### Remotely assessed short-term memory change is associated with long-term lPACC change

We found that changes in performance on a familiarity-dependent memory task (CSR) across 30 weeks were correlated with changes on the lPACC measured across eight years. This suggests that the CSR, when frequently administered over a relatively short time frame, captures change in a similar cognitive construct as established in-person neuropsychological tests designed to capture changes due to AD pathology. The MDT-O, measuring memory precision for objects, did not show a significant change-change relationship with lPACC score when looking at both groups, indicating that the longitudinal trajectories of the underlying constructs measured with these tests might be divergent. We were unable to investigate change-change correlations with the tasks of associative memory (ORR) or memory precision for scenes (MDT-S), as these tasks did not show sufficient variance in change, which is necessary to investigate covariances of change. This could be due to the short timeframe, meaning that changes in associative memory and memory precision for scenes happen over a longer period of time than only 30 weeks. Alternatively, this could be because the rate of change in those individuals with MCI had already slowed so as to be undetectable in the current sample.

### Associative memory and memory precision may be affected at earlier disease stages

Consistent with the hypothesis that decline across the different remote tasks would be sequential rather than simultaneous, we found that memory precision (MDT-OS) and associative memory (ORR) showed significant main effect of group, and that these baseline effects were descriptively larger than those seen for familiarity-dependent memory (CSR). Additionally, we did not observe differences in 30-week change between groups for the former tasks. We speculate that, while at an earlier point in time, there was decline in memory precision and associative memory such that a large group difference was observed at baseline, the rate of decline at the time of measurement was so slow as to be undetectable. That is, the peak rate of decline for memory precision and associative memory may occur during an earlier disease stage, before a clinical diagnosis of MCI is made. The observed difference in CSR change suggests that MCI might represent a sensitive period for familiarity-dependent memory, such that it is able to be captured within a 30-week window (see Fig. 5).

**Fig. 5.**
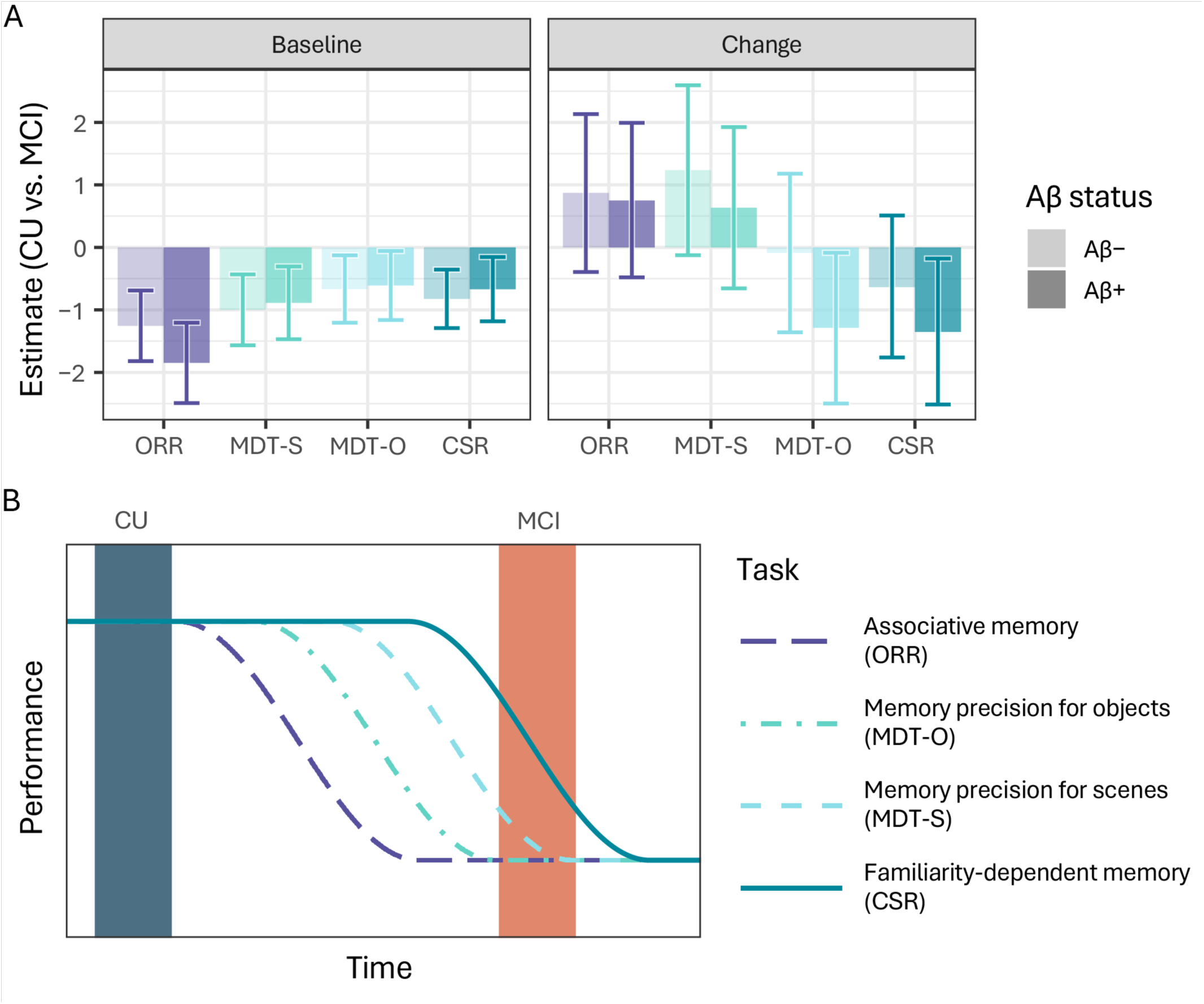
Decline in specific episodic memory processes may be sequential. (A) Estimates of group differences between cognitively unimpaired (CU) and mild cognitive impairment (MCI) groups stratified by Aβ status. Error bars represent 95% confidence intervals. (B) A hypothetical timeline of differential decline in task performance and the time windows of measurement in the current study in which associative memory measured by the Object-in-Room Recall (ORR) declines first, followed by the memory precision measured by the Mnemonic Discrimination Task for Scenes (MDT-S) and Objects (MDT-O), respectively, and with familiarity-dependent memory measured by the Complex Scene Recognition (CSR) performance declining last. The sensitive period for the ORR and MDT-S may be during an earlier disease stage, before MCI can be diagnosed.

This explanation is largely consistent with our hypotheses based on the spatiotemporal spread of tau pathology. We expected the MDT-OS and the ORR to show the earliest signs of decline along disease progression, due to their reliance on the hippocampal and entorhinal cortices, in which the earliest signs of pathological tau accumulation can be detected^18,19,64,65^. In the current sample, this manifested as more severe impairment in MCI at baseline and a slowed rate of decline. The CSR, in contrast, which relies on extrahippocampal familiarity-dependent memory^34^ and recruits a wide network of brain regions including the medial temporal lobe but also frontal and parietal regions^31–33^, which are typically affected by tau pathology later in course of disease progression, seems to have been relatively preserved until the MCI stage (i.e., prodromal AD), during which we observed decline. Information about tau burden was not available in the current sample, therefore our conclusions in this regard are speculative for now. Future research may directly investigate the effects of regional tau deposition on change in performance in these tasks, as well as declines in performance in preclinical AD samples, in which tau predominantly accumulates in the entorhinal cortex and hippocampus^37^.

Of note, the MCI group did not show floor effects with respect to possible task performance: mean corrected hit rate was 0.334 and 0.387 for the MDT-S and MDT-O, respectively (chance performance = 0), while mean ORR score was 17.7 (71%; chance performance = 33%). Additionally, only 3%, 2%, and 2% of the MCI group’s scores were below chance for the MDT-S, MDT-O, and CSR respectively, while no individual performed worse than chance on the ORR at any time point. This further suggests a disease-contingent asymptote (i.e., slowing of decline rate), rather than a psychometric floor effect caused by technical limitations of the tests.

### Limitations

There are a number of limitations that should be considered when interpreting the results of the current analyses. Firstly, this was a harmonized multi-cohort analysis in which the study designs across the three cohorts were not identical; in particular, in-person assessments differed across sites. While this was accounted for by deriving a latent score similar to the PACC, the underlying cognitive construct may have differed slightly across cohorts. Additionally, although the MDT-O and CSR were sensitive to the change in the current MCI sample, 30 weeks may not be a long enough period to see meaningful change in all aspects of episodic memory in early AD, when disease progression is still relatively slow; we may have seen stronger effects across longer time periods. Future studies may consider deploying remote and unsupervised cognitive tests over longer periods of time, and indeed, some studies with such designs are already planned or under way (e.g., REAL AD^66^, DELCODE 2). The MCI group included in the current analyses was also moderately sized, especially considering the stratification according to Aβ status. The generalization of the current results may therefore be limited until they can be replicated in other similar but larger samples.

Another limitation to the current study is that there was no regional tau information available as neither the DELCODE nor memory clinic cohorts underwent positron emission tomography (PET) scanning. We speculate about the mechanisms underlying the degradation of various episodic memory constructs across the AD spectrum based on earlier literature, but these hypotheses should be tested in samples in which all relevant information is available.

Finally, we hypothesize that associative memory as measured by the ORR and memory precision for scenes as measured by the MDT-S might detect decline in performance among individuals who are less advanced in the course of disease progression, such as those that are Aβ positive and might experience subjective cognitive decline but no objective cognitive impairment, that is, those with preclinical AD (i.e., clinical stage 2 according to recent NIA-AA criteria^15^). However, diagnostic information regarding SCD was not available for a large portion of the sample at the time of these analyses, therefore we were unable to look at decline during this early stage of AD.

### Implications for remote testing in clinical contexts

The results of the current study indicate that disease-relevant cognitive decline can be monitored during a relatively short timeframe using frequently administered remote and unsupervised testing. This has implications for the use of remote testing in clinical contexts, namely the use of individualized testing to collect additional information that may inform the diagnostic process, as well as to monitor disease progression and response to treatments. More research with large sample sizes will be required to develop norms in terms of cognitive decline on remote tasks. Once these norms are established, however, patients may be able to complete frequent remote testing using their own devices to measure their individual rate of cognitive decline. A clinician could then compare this to the expected rate of decline for that individual’s demographic group and determine whether they show conspicuous decline, facilitating the diagnostic process. This established rate of change could be used to evaluate the rate of change during a later time window in the case of individualized monitoring. The use of remote and unsupervised digital cognitive testing will increase the number of data points available to make important clinical decisions and will enable clinicians to better accompany their patients facing progressive memory loss over time.

## Conclusion

We found that (bi-)weekly remote and unsupervised cognitive assessments are feasible in older adults with and without cognitive impairment, showing that these cognitive tests are suitable for high-frequency assessment. This, in turn, improves their measurement reliability compared to yearly tests and thereby increasing their sensitivity to subtle cognitive decline. Indeed, remote tests of familiarity-dependent memory and memory precision for objects were sensitive to cognitive decline seemingly driven by AD pathology. Additionally, the patterns of change seen on the remote task of familiarity-dependent memory over only 30 weeks reflected a similar pattern of change captured by traditional neuropsychological assessments over multiple years. In-person neuropsychological test scores did not show group differences in short-term change, emphasizing the need for more sensitive assessment tools, for example, those that can be frequently administered. Finally, the remote tasks also showed differential patterns of change, suggesting that they may be sensitive to cognitive changes at different disease stages, though future studies are necessary to confirm this hypothesis. Taken together, the current findings support the use of smartphone- and tablet-based tasks to capture and monitor episodic memory decline in all-cause MCI and prodromal AD.

## Data Availability

All data produced in the present study are available upon reasonable request to the authors.

## Acknowledgements

We would like to thank all participants, patients, and caregivers, as well as stad members involved in DELCODE, WRAP, Wisconsin ADRC, and the Magdeburg memory clinic for their contributions to this study. Funding for this research has been provided by the Alzheimer Forschung Initiative e.V. (PI: David Berron), as well as NIA grants (R01 AG027161, P30 AG062715) and a UW-Madison Department of Medicine Pilot Program Grant (PI: Lindsay Clark). DELCODE is funded by Clinical Research, German Center for Neurodegenerative Diseases. We would also like to thank the project stad at neotiv for technological and project support. D.B. and E.D. are scientific co-founders of neotiv GmbH and own company shares.

