## Supplementary Materials for "Smartphone-based detection of memory decline in prodromal Alzheimer’s disease"

Polk et al. 2025

### Supplementary Methods

#### Study cohorts

Seventy-seven individuals recruited through the DZNE: Longitudinal Cognitive Impairment and Dementia Study (DELCODE)[1] were included in the current analyses. DELCODE is a multi-site observational study in Germany including extensive clinical and neuropsychological testing and magnetic resonance imaging (MRI), as well as blood, urine, and cerebrospinal fluid (CSF) sampling (see ref. [1] for study details). Individuals aged 60 years or older who had visited one of the ten memory clinics involved in DELCODE were recruited to participate in the longitudinal study with yearly assessments. Data collection started in April 2014 and is currently ongoing. At their baseline appointment, DELCODE participants were categorized as having healthy cognition, a subjective cognitive decline (SCD) diagnosis, or a mild cognitive impairment (MCI) diagnosis. An SCD diagnosis is given if the patient visited their general practitioner with memory complaints, but scored above −1.5 SD on the Consortium to Establish a Registry for Alzheimer's Disease (CERAD) neuropsychological test battery based on an age-, sex-, and education-adjusted normal performance range and fulfilled research criteria for SCD[2,3]. MCI diagnoses were made based on the current research criteria by the National Institute on Aging and Alzheimer’s Association (NIA-AA)[4,5]. Participants’ cognitive status was re-assessed at each follow-up appointment. DELCODE participants were asked at an annual in-person assessment if they would like to be included in the mobile add-on study. On average, participants had been enrolled in the parent study for 3.3 years (SD = 1.4) prior to starting the mobile add-on study, with the chronologically closest in-person assessment less than 0.1 years before the start of the mobile add-on study (SD = 0.1).

Another 35 individuals recruited through the memory clinic of the Department of Neurology and the Institute of Cognitive Neurology and Dementia Research at the Medical Faculty of the University Hospital of the Otto-von-Guericke University were included. These participants were also categorized as CU, SCD, or MCI using the same criteria as those used with DELCODE participants before the start of the mobile add-on study. On average, participants had their first memory clinic visit 0.9 years (SD = 1.1) prior to starting the mobile add-on study, with the closest in-person assessment less than 0.1 years after the start of the mobile add-on study (SD = 0.3).

Note that in the current analyses, participants from DELCODE and the memory clinic with healthy cognition and SCD are both considered cognitively unimpaired (CU), as they showed no clinical indication of cognitive impairment.

Finally, another 91 individuals recruited through the Wisconsin Registry for Alzheimer's Prevention (WRAP)[6,7] and the Wisconsin Alzheimer' Disease Research Center (ADRC) were included. WRAP and the Wisconsin ADRC are ongoing longitudinal observational cohort studies enriched for a family history of dementia of the Alzheimer's type at the University of Wisconsin-Madison. Together, participants recruited through WRAP and Wisconsin ADRC are referred to as the Wisconsin cohort, as the protocols related to the current analyses are identical. Study visits for the Wisconsin cohort are completed annually (Wisconsin ADRC participants with impairment or over 65 years old) or biennially (WRAP, Wisconsin ADRC participants 65 years old or younger) and include physical exams, neuropsychological testing, and questionnaires on medical and lifestyle histories. Additionally, CSF collection, [$^{11}$C]Pittsburgh compound B (PiB) positron emission tomographic (PET) imaging, and structural MRI are conducted in a subset of individuals. Study visit data are reviewed by a consensus conference panel comprising dementia-specialist physicians, neuropsychologists, and nurse practitioners, and participants classified as having healthy cognition, MCI, or dementia using standard diagnostic criteria[4,5]. Wisconsin participants who met inclusion criteria for the mobile add-on study were recruited over the phone. On average, participants had been enrolled in the parent study for 14.3 years (SD = 3.8) prior to starting the mobile add-on study, with the closest in-person assessment 0.4 years before the start of the mobile add-on study (SD = 1.1).

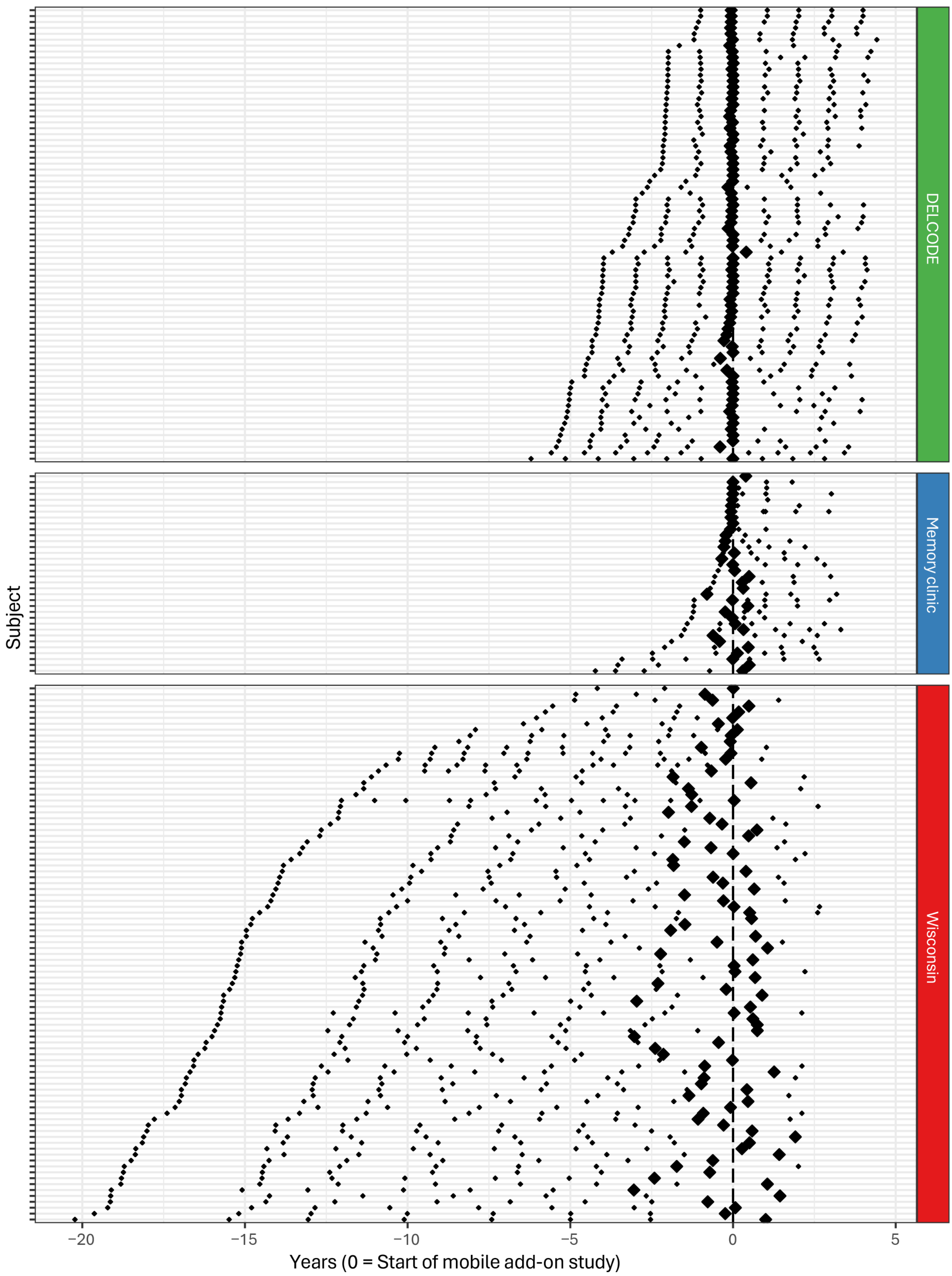


**Supplementary Fig. 1 | In-person assessments per subject.** Each black diamond indicates an in-person assessment through the parent study or at the memory clinic. Large black diamonds indicate the chronologically closest in-person appointment to the start of the mobile add-on study. In-person assessments from 4.5 years before and 4.5 years after the start of the mobild-add on study (0 on the x-axis) were considered in the current analyses.

**Amyloid-β acquisition**

For participants recruited through DELCODE, Aβ status was determined using $A\beta_{42}/A\beta_{40}$ ratio from lumbar CSF with a cut-off of ≤ 0.08 (Mesoscale Diagnostics LLC, Rockville, USA), with lower ratios indicating Aβ positivity, which was obtained through two-component Gaussian mixture modeling of $A\beta_{42}/A\beta_{40}$ ratios. If CSF data were not available for a participant, the individual probability of Aβ positivity in CSF was calculated using a binomial logistic regression model, which included age, number of APOE ε4 alleles [0/1/2], APOE ε2 carriership [yes/no], and plasma $A\beta_{42}/A\beta_{40}$ ratio (Lumipulse panel; Fujirebio Inc., Tokyo, Japan; see ref. [8]), following ref. [9]. A cut-off of > .639 was determined using the Youden index (cost ratio of false negative to false positive set to 1.5:1), with higher values indicating higher probability of CSF-based Aβ positivity. On average, Aβ measurements were taken 2.7 years before the start of the mobile add-on study (SD = 1.7, range = 6.2 years to 0.0 years before).

For participants recruited through the memory clinic, Aβ status was determined using CSF $A\beta_{42}/A\beta_{40}$ ratio with a cut-off of < 0.69 (Lumipulse panel; Fujirebio Inc., Tokyo, Japan), with lower values indicating Aβ positivity. On average, lumbar punctures were performed 0.3 years before the start of the mobile add-on study (SD = 0.8, range = 3.4 years before to 1.1 years after).

For the Wisconsin cohort, Aβ status was determined using the CSF $A\beta_{42}/A\beta_{40}$ ratio with a cut-off of ≤ .046 (NeuroToolKit panel; Roche Diagnostics International Ltd, Rotkreuz, Switzerland), with lower values indicating Aβ positivity. If CSF was not available for a participant, global cortical PiB with a cut-off of ≥ 1.19 distribution volume ratio (DVR) was used, with higher values indicating Aβ positivity. On average, Aβ measurements were taken 2.1 years before the start of the mobile add-on study (SD = 3.0, range = 12.0 years before to 1.4 years after).

**Neuropsychological testing**

Annual or biennal neuropscyhological testing was completed in German in the DELCODE and memory clinic cohorts and in English in the Wisconsin cohort. See Supplementary Table 1 and Supplementary Figs. 2 and 3 for details regarding the neuropsychological testing.

**Mini Mental State Examination (MMSE).** The MMSE[10] is a 30-item screening tool used to assess cognitive status. It was administered to all participants except those from the Wisconsin ADRC.

**Montreal Cognitive Assessment (MoCA).** The MoCA[11] is a 10-minute cognitive screening tool designed to help general practitioners detect MCI. It was administered in the Wisconsin ADRC and cross-walked with the MMSE, such that scores were on the same scale as the MMSE. The cross-walked MMSE (i.e., including rescaled MoCA scores) is included in the lPACC model.

**Category and phonemic fluency tests.** Three category/phonemic fluency tests were administered: groceries (DELCODE and memory clinic), words starting with a certain letter (memory clinic and Wisconsin), and animals (all cohorts). The Wisconsin cohort completed letter fluency for the letters C, F and L, while the memory clinic cohort only completed it for the letter S, therefore the Wisconsin cohort's scores were divided by three. These test semantic memory and executive function.

**Alzheimer's Disease Assessment Scale—Cognitive (ADAS-Cog) Word Recall.** The ADAS-Cog Word Recall subscale[12] is a 10-item word list to assess verbal recall. It was administered in the DELCODE and memory clinic cohorts.

**Free and Cued Selected Reminding Test (FCSRT96).** The FCSRT[13] is a test of free and cued recall of newly learned associations. The FCSRT96 is calculated as double the number of items freely recalled plus the number of items recalled with a cue and ranges from 0 to 96. This test was administered in the DELCODE and memory clinic cohorts.

**Rey Auditory Verbal Learning Test (RAVLT).** The RAVLT[14] is a 15-item word list to assess verbal recall. It was administered in the Wisconsin cohort.

**Symbol Digit Modalities Test (SDMT).** The SDMT[15] is a 90-item test measuring associative memory, executive function, and processing speed. It was administered in the DELCODE and memory clinic cohorts.

**Trail Making Test B (TMT B).** The TMT B tests sustained attention and task switching[16]. It was administered in all cohorts.

**Digit Symbol Test (DST)**. The DST[17] is a 93-item test from the Wechsler Adult Intelligence Scale—Revised (WAIS-R) measuring associative memory, executive function, and processing speed. It was administered in the Wisconsin cohort.

**Logical Memory Delayed Recall (LMDR).** The Logical Memory subscale from the Wechsler Memory Scale (WMS) tests story recall after a 30-minute delay. Version A from the Revised version (WMS-R)[18] was administered in WRAP, and version B from the Fourth version (WMS-IV)[19] was administered in the DELCODE and memory clinic cohorts.

**Craft Story 21.** The Craft Story 21[20] is a test of story recall. It was administered in the Wisconsin ADRC and cross-walked with the LMDR A, such that scores were on the same scale as the LMDR. The cross-walked LMDR A (i.e., including rescaled Craft Story 21 scores) is included in the lPACC model.

**Supplementary Table 1.**

*Information about in-person measurements, including means and standard deviations of in-person neuropsychological tests per cohort at the in-person appointment chronologically closest to the start of the mobile add-on study, as well as loading obtained in the first step of the lPACC estimation.*

| | CU | MCI | DELCODE | Memory clinic | Wisconsin |
|---|---|---|---|---|---|
| Time points included per person (M ± SD) | 4.2 ± 2.3 | 4.5 ± 1.9 | 6.6 ± 1.3 | 3.9 ± 1.4 | 2.6 ± 1.0 |
| Time range of tests included (years) | −4.5 to 4.2 | −4.3 to 4.4 | −4.5 to 4.4 | −4.2 to 3.3 | −4.5 to 2.7 |
| Chronologically closest in-person appointment (years) | −0.24 ± 0.85 | −0.05 ± 0.37 | −0.05 ± 0.10 | 0.01 ± 0.30 | −0.39 ± 1.09 |

| Task | Obs. | CU | MCI | DELCODE | Memory clinic | Wisconsin | λ |
|---|---|---|---|---|---|---|---|
| MMSE/MoCA | 838 | 29.5 ± 0.8 | 26.1 ± 4.0 | 29.2 ± 1.3 | 24.5 ± 4.4 | 29.5 ± 0.8 | .901 |
| Fluency: Groceries | 532 | 25.3 ± 5.2 | 19.0 ± 4.9 | 24.3 ± 5.5 | 15.6 ± 5.8 | n.a. | .771 |
| Fluency: Phonemic | 337 | 15.5 ± 4.0 | 9.7 ± 4.5 | *n.a.* | 10.1 ± 4.9 | 15.5 ± 4.0 | .704 |
| Fluency: Animals | 863 | 25.0 ± 5.8 | 17.9 ± 5.7 | 24.8 ± 6.0 | 16.7 ± 6.4 | 23.1 ± 5.5 | .637 |
| ADAS-Cog Word Recall | 594 | 8.4 ± 1.6 | 4.0 ± 2.7 | 7.7 ± 2.3 | 3.4 ± 2.7 | *n.a.* | .919 |
| FCSRT96 | 457 | 81.8 ± 5.9 | 67.5 ± 15.0 | 79.5 ± 8.0 | 58.2 ± 24.5 | *n.a.* | .882 |
| RAVLT | 236 | 52.0 ± 8.5 | 29.2 ± 3.7 | *n.a.* | *n.a.* | 51.4 ± 9.2 | .819 |
| SDMT | 527 | 51.7 ± 9.7 | 39.9 ± 12.1 | 49.7 ± 10.4 | 32.2 ± 15.8 | *n.a.* | .864 |
| TMT B | 769 | 74.5 ± 30.6 | 131.5 ± 64.1 | 91.6 ± 42.7 | 133.0 ± 66.4 | 59.9 ± 28.2 | −.833 |
| DST | 205 | 55.9 ± 10.4 | 37.8 ± 9.9 | *n.a.* | *n.a.* | 55.3 ± 10.8 | .926 |
| LMDR A/Craft Story 21 | 233 | 13.7 ± 3.7 | 5.5 ± 3.3 | *n.a.* | *n.a.* | 13.5 ± 3.9 | .635 |
| LMDR B | 534 | 16.0 ± 4.1 | 10.2 ± 5.6 | 15.2 ± 4.5 | 5.1 ± 5.3 | *n.a.* | .908 |

*Note.* MMSE = Mini-Mental State Examination, ADAS-Cog = Alzheimer's Disease Assessment Scale—Cognitive; FCSRT = Free and Cued Selected Reminding Test, RAVLT = Rey Auditory Verbal Learning Test, SDMT = Symbol Digit Modalities Test, TMT B = Trail Making Test B, DST = Digit Symbol Test, LMDR = Logical Memory Delayed Recall, CU = cognitively unimpaired, MCI = mild cognitive impairment, *n.a.* = not administered, λ = loading onto the latent Preclinical Alzheimer's Cognitive Composite (lPACC).

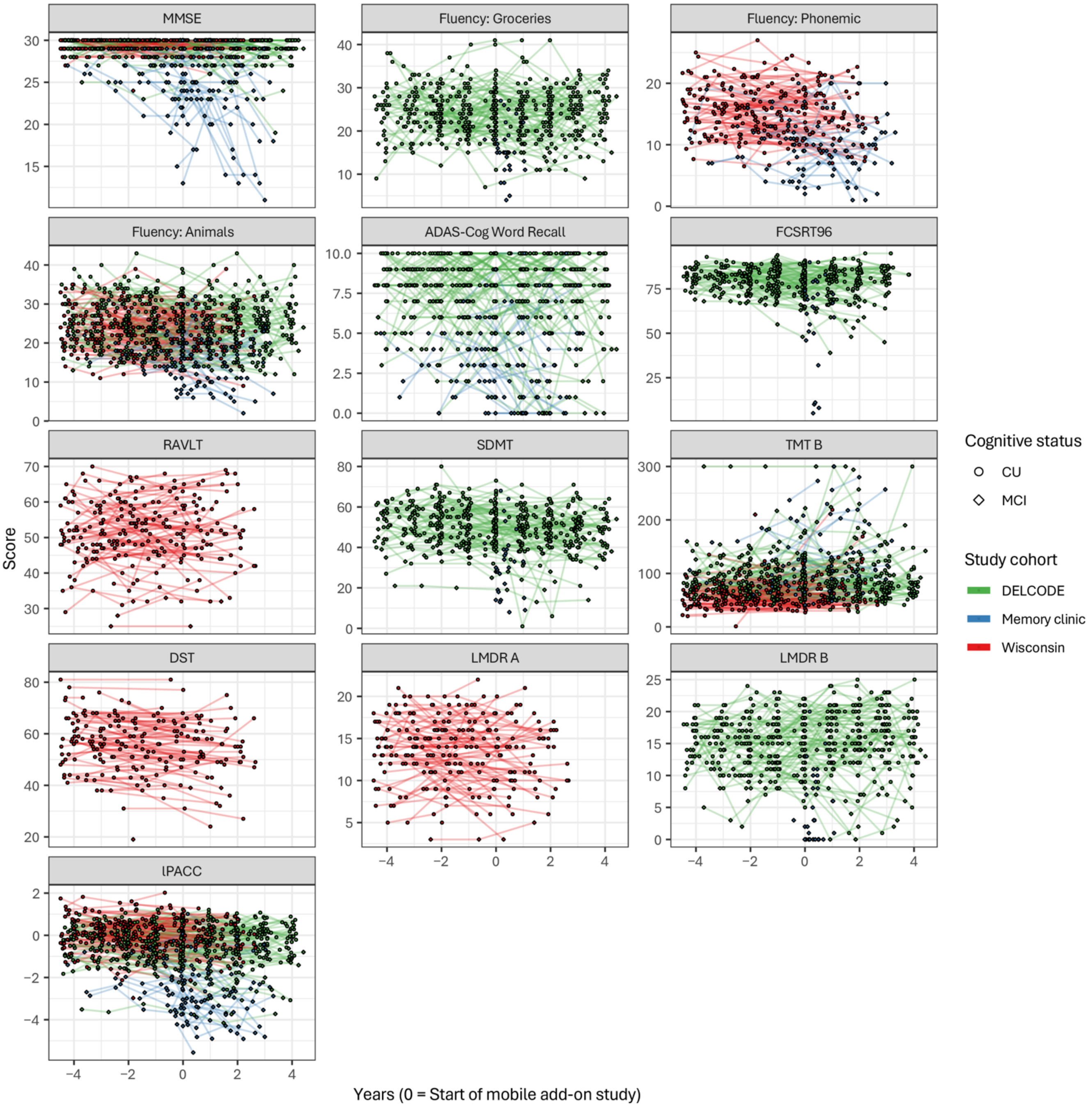


**Supplementary Fig. 2 | Raw scores of the in-person neuropsychological assessments as well as the extracted lPACC scores.** MMSE = Mini-Mental State Examination, ADAS-Cog = Alzheimer's Disease Assessment Scale—Cognitive; FCSRT = Free and Cued Selected Reminding Test, RAVLT = Rey Auditory Verbal Learning Test, SDMT = Symbol Digit Modalities Test, TMT B = Trail Making Test B, DST = Digit Symbol Test, LMDR = Logical Memory Delayed Recall, lPACC = latent Preclinical Alzheimer's Cognitive Composite, CU = cognitively unimpaired, MCI = mild cognitive impairment.

***Estimation of lPACC scores with confirmatory factor analysis***

To harmonize neuropsychological test scores across cohorts, a latent score was derived for each individual using structural equation modeling with the *lavaan* package in R[21]. In a first step, a confirmatory factor analysis (CFA) model was built using the scores from the chronologically closest in-person appointment from each participant as manifest variables, scaled such that the CU group had a mean of 0 and a standard deviation of 1. Each of the manifest variables loaded onto a latent factor, which we call latent Preclinical Alzheimer's Cognitive Composite (lPACC), and each of the loadings was freely estimated. To ensure that the model was fully identifiable, the variance of lPACC was fixed to 1. The residual variance of each manifest variable was freely estimated with a lower bound of .001. Residual covariances between the fluency variables were also freely estimated because of the similarity between the tasks. The model was run using maximum likelihood estimation with robust (Huber-White) standard errors (MLR). Missing data were handled using maximum likelihood estimation (ML). The standardized loadings of each manifest variable onto lPACC from the first step were saved (see Supplementary Table 1). This model showed acceptable fit according to ref. [22], CFI = .912, RMSEA = .069. In a second step, all data from 4.5 years before to 4.5 years after the start of the mobile add-on study were included in a CFA. The loadings of the manifest variables onto lPACC were fixed to the loadings saved from the first step. The variance of lPACC was freely estimated with a lower bound of .001and the mean was fixed to 1. Residual variances were freely estimated with a lower bound of .001. The model was run using MLR and missing data were handled with ML. Each subject's lPACC score at each time point was extracted and used in subsequent analyses.

### Mobile add-on study

Participants were reminded to complete the tasks via push notifications; they could postpone task completion if the notification came at an inconvenient time (e.g., the participant was currently distracted, fatigued, ill, etc.). Each task was performed in two phases with a delay between phases (30 minutes to 24 hours; see below for details), and the second phase was also initiated via push notification. Before participants started the tasks, they were requested to perform the task in a quiet environment, to put on glasses if necessary, and to adjust the settings of their device so they could clearly see the task stimuli. After each test session, participants answered a short questionnaire within the app regarding distraction during the task (yes/no), as well as concentration level and subjective performance (1 = very bad, 2 = bad, 3 = neither good nor bad, 4 = good, 5 = very good) during the task. All task instructions were administered remotely through the app (in German for DELCODE and memory clinic participants, in English for Wisconsin participants) and participants performed all tasks fully unsupervised. See Fig. 1 in the main text for examples of how each task looked in the app.

**Object-in-Room Recall (ORR).** In the ORR, which tested associative memory, participants saw a series of 25 computer-generated rooms rendered in 3D; two 3D-rendered objects were presented in each room. After each stimulus was presented, participants saw a location indicated by a colored circle in the empty room and were instructed to indicate which of three objects (target, lure in the correct room but wrong location, or unfamiliar lure) they had seen in that specific location (immediate recall). After a 30-minute (DELCODE at odd-numbered sessions and memory clinic), 90-minute (Wisconsin), or a 24-hour delay (DELCODE at even-numbered sessions), participants were notified that the second phase (delayed recall) could be completed. The delayed recall phase was identical to the immediate recall phase. The median time it took to complete both the learning and retrieval phases was 13.9 minutes. The average score between immediate and delayed recall was calculated and used in subsequent analyses, and time elapsed between the immediate and delayed recall phases was taken into account in the models as a nuisance variable. Wisconsin participants completed the ORR task first.

**Mnemonic Discrimination Task for Objects and Scenes (MDT-OS).** In the MDT-OS, which tested memory precision for object and scene stimuli, participants saw computer generated objects and scenes rendered in 3D followed by either an identical repeat image or a similar but not identical lure image. Participants were instructed to indicate via button press if the current image was identical to the one seen before, or to tap on the part of the image that had changed. Thirty-two object pairs and 32 scene pairs were presented, with half of the pairs containing repeated stimuli and half containing similar but not identical stimuli. During the first phase (one-back), one pair of images was presented in each trial. After a 24-hour (DELCODE, memory clinic, and Wisconsin burst design) or two-week delay (Wisconsin continuous design), participants were notified that the second phase (two-back) could be completed. Note that the second phase was not dependent on the first, thus we assume that the length of the delay between the two phases has a negligible effect on performance and do not correct for this. During the two-back phase, two interleaved pairs of images were presented in each trial. The median time it took to complete both phases of the MDT was 19.9 minutes. The corrected hit rate, that is, the hit rate (percentage of correctly identified repeat items) minus the false alarm rate (percentage of incorrectly identified lure items), was calculated for both phases for objects and scenes separately, and the average corrected hit rate between phases was used in subsequent analyses. The MDT-O and MDT-S were analyzed separately.

**Complex Scene Recognition (CSR).** In the CSR, which tested familiarity-dependent memory, participants saw 60 photographic images depicting scenes. During the first phase (encoding), participants were instructed to indicate via button press whether each scene was indoor or outdoor. After a 65-minute delay, participants were notified that the second phase (recognition) could be completed. During the recognition phase, the encoded images, along with 30 new images, were presented to participants. They were instructed to indicate whether they had seen the current image before or not, or if they were unsure. The median time it took to complete both learning and retrieval phases was 13.2 minutes. The corrected hit rate was used in subsequent analyses, and time elapsed between the learning and recognition phases was taken into account. DELCODE and memory clinic participants completed the CSR as their first task.

***Data filtering***

Remote task data were filtered according to a number of criteria to ensure data quality. First, those ORR and CSR trials during which more than 96 hours elapsed before the retrieval phase, as were those trials for which the retrieval phase was missing. MDT-OS trials for which the 2-back phase was missing were likewise excluded. Trials with too many timed-out items were excluded (> 8 for ORR during either encoding or retrieval, > 17 for MDT-OS in both phases, > 13 for CSR during encoding and > 19 during retrieval). Trials during which technical errors (the majority of which were randomization errors) occurred were excluded. And finally, participants with only one observation for a given task were excluded. Additionally, due to a technical error, one participant completed 18 sessions before being reset and then completed another 24 sessions; only the data from this individual's first 18 sessions are considered. See Supplementary Table 2 for specifics regarding filtering. Supplementary Fig. 3 shows pairwise correlations between task performance at baseline after filtering, and Supplementary Fig. 4 shows the raw scores longitudinally and illustrates that there are minimal floor and ceiling effects. Finally, Supplementary Fig. 5 shows baseline correlations between demographic factors, in-person test scores, and remote test scores at baseline.i

**Supplementary Table 2.**

Number of observations filtered from each task before analyses.

| Task | $n_{CU}/n_{MCI}$ | Total obs. | Reason for filtering | Obs. (%) filtered | Obs. (%) included |
|---|---|---|---|---|---|
| ORR | 145/49 | 1153 | Time elapsed before retrieval phase > 96 hours | 53 (5%) | 1023 (89%) |
| | | | Missing retrieval phase | 16 (1%) | |
| | | | > 8 timeouts during encoding | 7 (1%) | |
| | | | > 8 timeouts during retrieval | 44 (4%) | |
| | | | Technical error | 2 (< 1%) | |
| | | | Only one observation per participant | 8 (1%) | |
| MDT-OS | 137/46 | 1102 | > 17 timeouts during task | 1 (< 1%) | 1003 (91%) |
| | | | Technical error | 91 (8%) | |
| | | | Only one observation per participant | 7 (1%) | |
| CSR | 148/49 | 1139 | Time elapsed before retrieval phase > 96 hours | 46 (4%) | 1067 (94%) |
| | | | Missing retrieval phase | 2 (< 1%) | |
| | | | > 13 timeouts during encoding | 11 (1%) | |
| | | | Technical error | 2 (< 1%) | |
| | | | Only one observation per participant | 11 (1%) | |

*Note.* CU = cognitively unimpaired, MCI = mild cognitive impairment, obs. = observations, ORR = Object-in-Room Recall, MDT-OS = Mnemonic Discrimination Task for Objects and Scenes, CSR = Complex Scene Recognition.

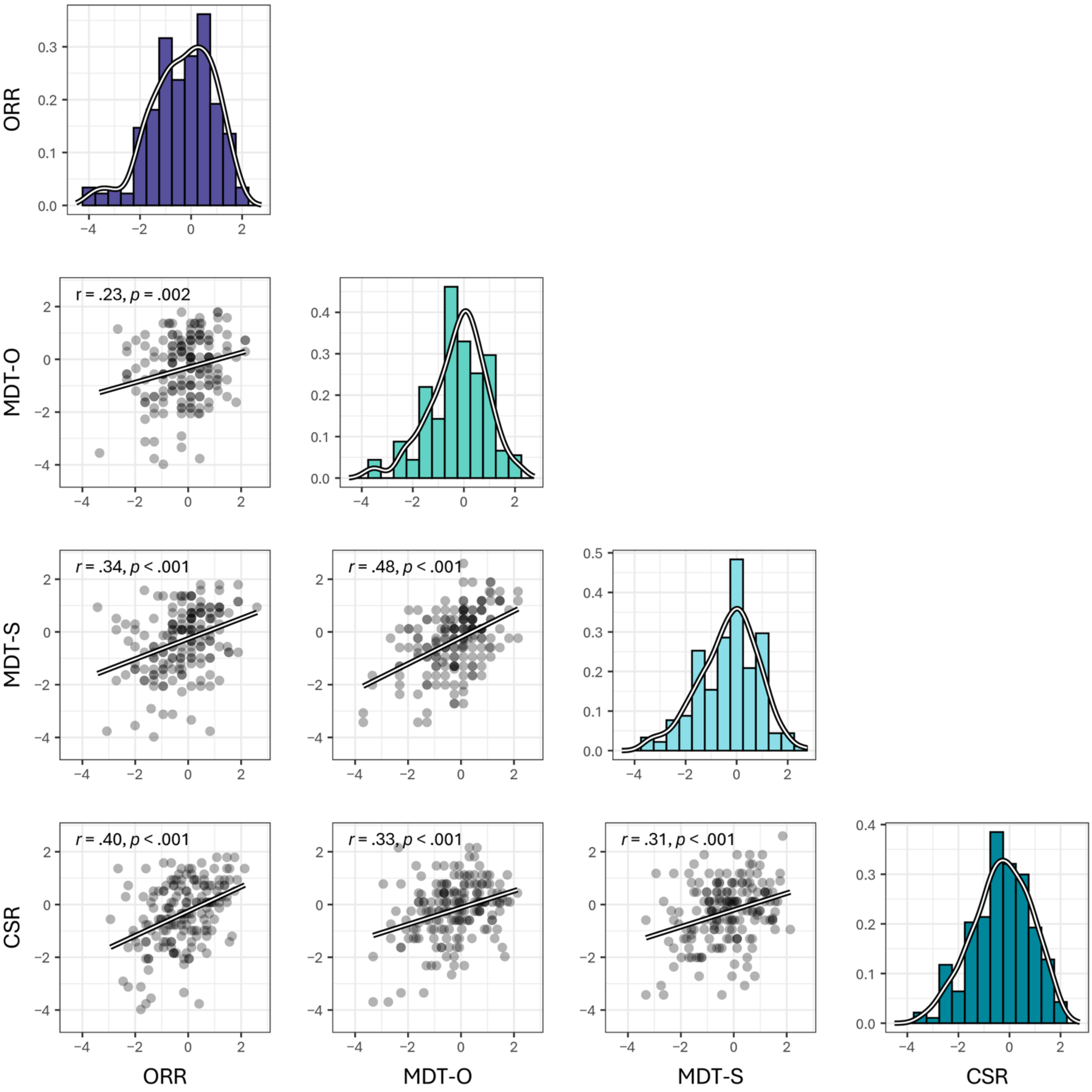


**Supplementary Fig. 3 | Pairwise correlations between performance on remote tasks at baseline.** Density plots showing the distribution of task performance (centered to the CU group and scaled) is shown on the diagonal. Pairwise scatterplots are shown in the lower triangle.
ORR = Object-in-Room Recall, MDT-O/-S = Mnemonic Discrimination Task for Objects/Scenes, CSR = Complex Scene Recognition.

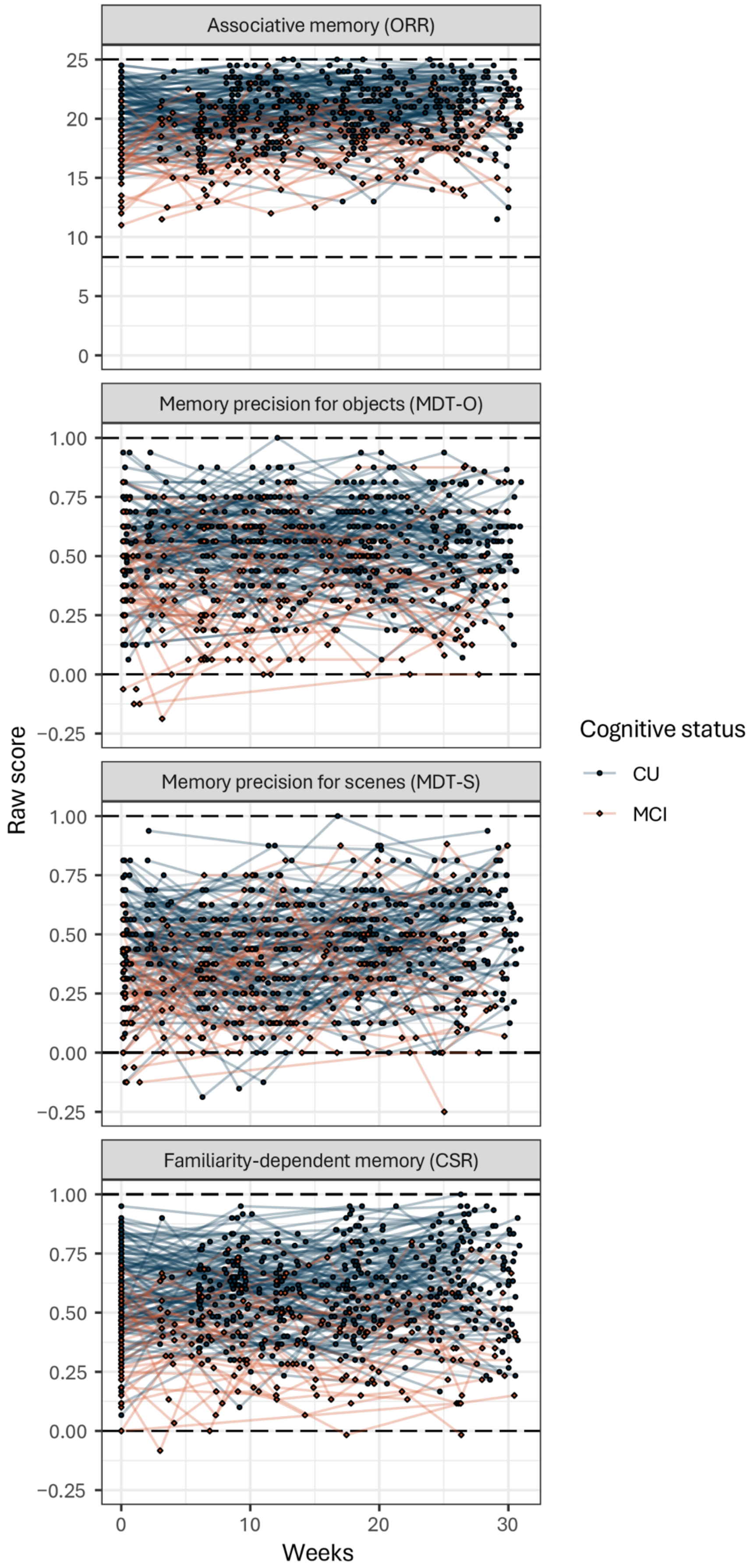


**Supplementary Fig. 4 | Raw scores on remote tasks over time.** Spaghetti plots showing individual performance over time on the remote tasks. For the ORR, average score between immediate and delayed recall is shown. For the MDT-O/-S, as well as for the CSR, corrected hit rates (hit rate – false alarm rate) are shown. Higher scores indicate better performance on all tasks. Dashed lines represent chance performance (i.e., floor) and the highest score possible (i.e., ceiling).
ORR = Object-in-Room Recall; MDT-O/-S = Mnemonic Discrimination Task for Objects/Scenes; CSR = Complex Scene Recognition.

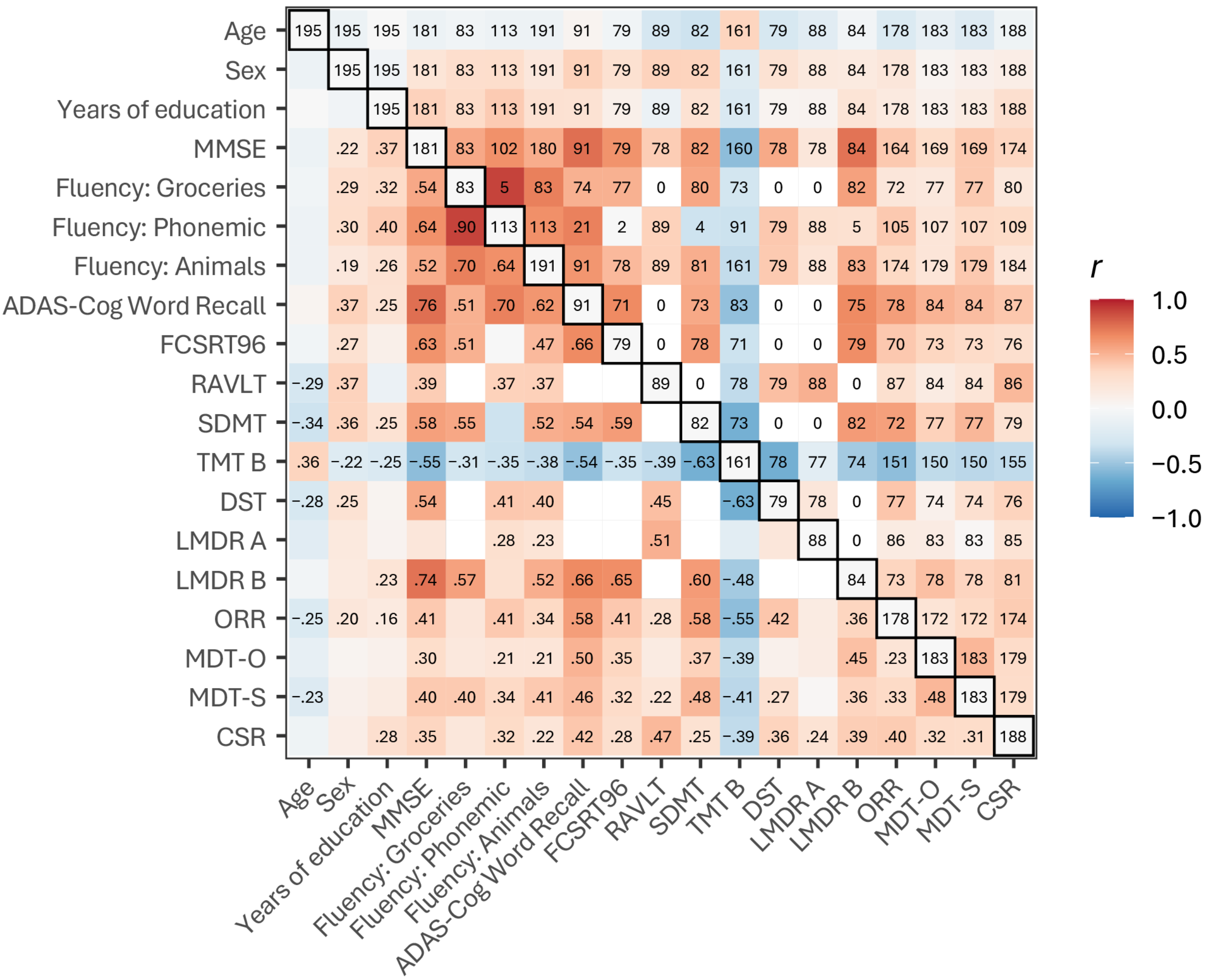

**Supplementary Fig. 5 | Pairwise correlations between demographic factors, neuropsychological test scores at the in-person visit chronologically closest to the start of the mobile add-on study, and remote task scores at baseline.** Correlation strength is indicated by color, with red indicating a positive correlation and blue indicating a negative correlation. The *N* of participants included in each pair is given on the diagonal. The *n*s included in each pairwise correlation are found in the upper triangle. Correlation coefficients *r* of significant pair-wise correlations at $p < .050$ uncorrected are found in lower triangle. Note that some pairs of tests have no overlap (white cells) as they were not administered in the same cohorts. Sex is coded as 0 for male and 1 for female.
MMSE = Mini-Mental State Examination, ADAS-Cog = Alzheimer's Disease Assessment Scale—Cognitive; FCSRT = Free and Cued Selected Reminding Test, RAVLT = Rey Auditory Verbal Learning Test, SDMT = Symbol Digit Modalities Test, TMT B = Trail Making Test B, DST = Digit Symbol Test, LMDR = Logical Memory Delayed Recall, ORR = Object-in-Room Recall, MDT-OS = Mnemonic Discrimination Task for Objects and Scenes, CSR = Complex Scene Recognition.

***Calculation of adherence and compliance to the mobile add-on study***

To calculate adherence, the number of task sessions completed by an individual was divided by the maximum number of task session possible for their cohort, which was as follows:

- DELCODE: 24 task sessions
- Memory clinic: 30 task sessions
- Wisconsin: 18 task sessions

For example, completing 15 task sessions would result in an adherence rate of 63% for DELCODE partcipants, 50% for memory clinic participants, and 83% for Wisconsin participants.

To calculate compliance, first, the time spent in the study was calculated for each individual based on the last task they completed. Then, the possible number of task sessions that could have been completed during this time frame was calculated based on the interval of testing, which was as follows:

- DELCODE: One task session every 14 days (i.e., every other week)
- Memory clinic: One task session every 7 days (i.e., once a week)
- Wisconsin: One task session every 20 days (i.e., three task sessions every two months)

For example, an individual in DELCODE whose last task session was completed on day 100 could have completed 8 tasks in this time frame (on days 0, 14, 28, 42, 56, 70, 84, and 98). If they completed 6 tasks in this time, they would have a compliance rate of 6 / 8 = 75%. An individual from the memory clinic whose last task session was completed on day 100 could have completed 15 tasks (on days 0, 7, 14, 21, 28, 35, 42, 49, 56, 63, 70, 77, 84, 91, and 98). If they completed 6 tasks in this time, they would have a compliance rate of 6 / 15 = 40%. Finally, if an individual from the Wisconsin cohort whose last task session was completed on day 100 could have completed 6 tasks (on days 0, 20, 40, 60, 80, and 100). If they completed 6 tasks in this time, they would have a compliance rate of 6 / 6 = 100%.

### Supplementary Results

#### Supplementary Table 3.

Linear mixed model results for the lPACC comparing CU and MCI groups.

| Parameter | Unstandardized estimate (*b*) | Standardized estimate (*β*) | *p*-value |
|---|---|---|---|
| Intercept | −0.382 [−0.582, −0.182] | 0.184 [0.029, 0.338] | **< .001** |
| **Group [MCI]** | −1.455 [−1.774, −1.138] | −1.096 [−1.341, −0.853] | **< .001** |
| **Years** | −0.019 [−0.039, 0.000] | −0.033 [−0.068, 0.001] | .057 |
| **Years * Group [MCI]** | −0.145 [−0.189, −0.102] | −0.250 [−0.326, −0.176] | **< .001** |
| Age | −0.149 [−0.248, −0.051] | −0.116 [−0.192, −0.040] | **.003** |
| Sex [female] | 0.533 [0.340, 0.725] | 0.413 [0.264, 0.563] | **< .001** |
| Years of education | 0.220 [0.118, 0.323] | 0.171 [0.092, 0.250] | **< .001** |
| Study cohort [Memory clinic] | −0.856 [−1.216, −0.493] | −0.665 [−0.944, −0.382] | **< .001** |
| Study cohort [Wisconsin] | −0.100 [−0.319, 0.119] | −0.077 [−0.248, 0.093] | .366 |

*N* = 191, 861 observations included in the model. Group [CU], Years * Group [CU], Sex [male], and Study cohort [DELCODE] serve as reference points.

| | | Group-wise estimates (*b*) |
|---|---|---|
| Group | CU | −0.471 [−0.637, −0.305] |
| | MCI | −1.926 [−2.154, −1.698] |
| Years * Group | CU | −0.019 [−0.039, 0.001] |
| | MCI | −0.164 [−0.203, −0.125] |

*Note.* Parameters in bold are discussed in the main text. *P*-values in bold are significant at $p < .050$ uncorrected. CU = cognitively unimpaired, MCI = mild cognitive impairment.

**Supplementary Table 4.**

Linear mixed model results for the lPACC comparing CU, MCI Aβ− and MCI Aβ+ groups.

| Parameter | Unstandardized estimate (*b*) | Standardized estimate (*β*) | *p*-value |
|---|---|---|---|
| Intercept | −0.352 [−0.545, −0.160] | 0.192 [0.042, 0.342] | < **.001** |
| **Group [MCI Aβ−]** | −1.120 [−1.466, −0.775] | −0.865 [−1.133, −0.598] | < **.001** |
| **Group [MCI Aβ+]** | −1.891 [−2.271, −1.514] | −1.423 [−1.717, −1.130] | < **.001** |
| **Years** | −0.018 [−0.037, −0.001] | −0.032 [−0.064, −0.001] | **.042** |
| **Years * Group [MCI Aβ−]** | −0.048 [−0.103, 0.005] | −0.085 [−0.179, 0.008] | .075 |
| **Years * Group [MCI Aβ+]** | −0.246 [−0.302, −0.193] | −0.430 [−0.527, −0.336] | < **.001** |
| Age | −0.151 [−0.248, −0.055] | −0.118 [−0.194, −0.043] | **.002** |
| Sex [female] | 0.499 [0.309, 0.688] | 0.391 [0.242, 0.539] | < **.001** |
| Years of education | 0.217 [0.116, 0.319] | 0.170 [0.091, 0.249] | < **.001** |
| Study cohort [Memory clinic] | −0.869 [−1.229, −0.506] | −0.680 [−0.962, −0.396] | < **.001** |
| Study cohort [Wisconsin] | −0.111 [−0.327, 0.104] | −0.087 [−0.256, 0.082] | .304 |

*N* = 187, 846 observations included in the model. Group [CU], Years * Group [CU], Sex [male], and Study cohort [DELCODE] serve as reference points.

| | | Group-wise estimates (*b*) | Difference from MCI Aβ− |
|---|---|---|---|
| Group | CU | −0.463 [−0.624, −0.301] | |
| | MCI Aβ− | −1.583 [−1.877, −1.288] | |
| | MCI Aβ+ | −2.354 [−2.657, −2.051] | −0.771 [−1.256, −0.286] |
| Years * Group | CU | −0.018 [−0.036, 0.000] | |
| | MCI Aβ− | −0.067 [−0.118, −0.016] | |
| | MCI Aβ+ | −0.265 [−0.317, −0.213] | −0.198 [−0.285, −0.111] |

*Note.* Parameters in bold are discussed in the main text. *P*-values in bold are significant at $p < .050$ uncorrected. CU = cognitively unimpaired, MCI Aβ− = mild cognitive impairment without amyloid pathology, MCI Aβ+ = mild cognitive impairment with amyloid pathology.

**Supplementary Table 5.**

Linear mixed model results for the MDT-O comparing CU and MCI groups.

| Parameter | Unstandardized estimate (*b*) | Standardized estimate (*β*) | *p*-value | $\eta^2_p$ |
|---|---|---|---|---|
| Intercept | 0.192 [−0.131, 0.515] | 0.133 [−0.234, 0.502] | .244 | |
| **Group [MCI]** | −0.587 [−1.056, −0.118] | −0.733 [−1.076, −0.393] | **.014** | 0.029 |
| **Years** | −0.964 [−1.994, 0.065] | −0.158 [−0.326, 0.011] | .065 | |
| **Years * Group [MCI]** | −0.990 [−2.069, 0.086] | −0.162 [−0.338, 0.014] | .072 | |
| Age | −0.219 [−0.343, −0.095] | −0.192 [−0.300, −0.083] | **.001** | 0.064 |
| Sex [female] | −0.001 [−0.248, 0.245] | −0.001 [−0.217, 0.215] | .995 | 0.000 |
| Years of education | −0.024 [−0.151, 0.102] | −0.021 [−0.132, 0.089] | .701 | 0.001 |
| Study cohort [Memory clinic] | −0.510 [−0.976, −0.042] | −0.447 [−0.856, −0.037] | **.033** | 0.023 |
| Study cohort [Wisconsin] | −0.016 [−0.298, 0.265] | −0.014 [−0.261, 0.232] | .913 | |
| Concentration | −0.001 [−0.078, 0.077] | 0.000 [−0.069, 0.068] | .989 | 0.000 |
| Distraction [yes] | −0.104 [−0.344, 0.134] | −0.092 [−0.301, 0.118] | .390 | 0.001 |
| Time of day [12PM to 6PM] | −0.126 [−0.291, 0.038] | −0.111 [−0.255, 0.033] | .132 | |
| Time of day [6PM to 12AM] | −0.247 [−0.436, −0.058] | −0.216 [−0.382, −0.051] | **.011** | 0.012 |
| Time of day [12AM to 6AM] | −0.432 [−0.961, 0.095] | −0.379 [−0.843, 0.083] | .107 | |
| Task session [2] | 0.184 [−0.053, 0.420] | 0.161 [−0.046, 0.368] | .127 | |
| Task session [3] | 0.297 [−0.062, 0.656] | 0.260 [−0.054, 0.575] | .103 | |
| Task session [4] | 0.471 [−0.007, 0.948] | 0.413 [−0.006, 0.831] | .053 | |
| Task session [5] | 0.642 [0.072, 1.212] | 0.563 [0.063, 1.062] | **.027** | |
| Task session [6] | 0.991 [0.239, 1.743] | 0.869 [0.210, 1.528] | **.010** | 0.020 |
| Task session [7] | 0.902 [0.079, 1.724] | 0.790 [0.069, 1.511] | **.031** | |
| Task session [8] | 1.210 [0.291, 2.128] | 1.061 [0.255, 1.866] | **.009** | |
| Task session [9] | 0.956 [−0.067, 1.984] | 0.838 [−0.059, 1.739] | .065 | |
| Task session [10] | 1.396 [0.235, 2.560] | 1.224 [0.206, 2.244] | **.018** | |

*N* = 174, 717 observations included in the model. Group [CU], Years * Group [CU], Sex [male], Study cohort [DELCODE], Distraction [no], Time of day [6AM to 12PM], and Task session [1] serve as reference points.

| | | Group-wise estimates (*b*) |
|---|---|---|
| Group | CU | 0.487 [−0.066, 1.040] |
| | MCI | −0.100 [−0.826, 0.626] |
| Years * Group | CU | −0.964 [−2.014, 0.086] |
| | MCI | −1.954 [−3.575, −0.333] |

*Note.* Parameters in bold are discussed in the main text. *P*-values in bold are significant at $p < .050$ uncorrected. CU = cognitively unimpaired, MCI = mild cognitive impairment.

**Supplementary Table 6.**

Linear mixed model results for the MDT-O comparing CU, MCI Aβ− and MCI Aβ+ groups.

| Parameter | Unstandardized estimate (*b*) | Standardized estimate (*β*) | *p*-value | $\eta^2_p$ |
|---|---|---|---|---|
| Intercept | 0.191 [−0.131, 0.513] | 0.064 [−0.312, 0.441] | .244 | |
| **Group [MCI Aβ−]** | −0.667 [−1.205, −0.127] | −0.609 [−1.014, −0.206] | **.016** | 0.046 |
| **Group [MCI Aβ+]** | −0.608 [−1.162, −0.057] | −0.825 [−1.237, −0.415] | **.031** | |
| **Years** | −1.115 [−2.148, −0.082] | −0.184 [−0.355, −0.014] | **.034** | 0.016 |
| **Years * Group [MCI Aβ−]** | −0.088 [−1.360, 1.180] | −0.015 [−0.225, 0.195] | .891 | 0.033 |
| **Years * Group [MCI Aβ+]** | −1.287 [−2.499, −0.087] | −0.212 [−0.413, −0.014] | **.037** | |
| Age | −0.232 [−0.357, −0.107] | −0.205 [−0.315, −0.094] | **< .001** | 0.070 |
| Sex [female] | −0.006 [−0.256, 0.244] | −0.005 [−0.227, 0.215] | .965 | 0.000 |
| Years of education | −0.009 [−0.137, 0.119] | −0.008 [−0.121, 0.106] | .893 | 0.000 |
| Study cohort [Memory clinic] | −0.566 [−1.039, −0.092] | −0.500 [−0.918, −0.081] | **.019** | 0.028 |
| Study cohort [Wisconsin] | −0.015 [−0.298, 0.267] | −0.013 [−0.263, 0.236] | .916 | |
| Concentration | 0.003 [−0.076, 0.081] | 0.002 [−0.067, 0.072] | .950 | 0.000 |
| Distraction [yes] | −0.080 [−0.322, 0.162] | −0.070 [−0.285, 0.143] | .517 | 0.001 |
| Time of day [12PM to 6PM] | −0.126 [−0.292, 0.040] | −0.111 [−0.258, 0.035] | .135 | |
| Time of day [6PM to 12AM] | −0.242 [−0.434, −0.050] | −0.214 [−0.383, −0.044] | **.014** | 0.012 |
| Time of day [12AM to 6AM] | −0.450 [−0.975, 0.074] | −0.397 [−0.861, 0.065] | .090 | |
| Task session [2] | 0.210 [−0.035, 0.454] | 0.185 [−0.031, 0.401] | .092 | |
| Task session [3] | 0.352 [−0.018, 0.721] | 0.311 [−0.016, 0.637] | .062 | |
| Task session [4] | 0.537 [0.047, 1.027] | 0.475 [0.041, 0.907] | **.032** | |
| Task session [5] | 0.702 [0.123, 1.282] | 0.620 [0.109, 1.133] | **.017** | |
| Task session [6] | 1.254 [0.483, 2.026] | 1.108 [0.427, 1.790] | **.001** | 0.040 |
| Task session [7] | 1.267 [0.429, 2.107] | 1.119 [0.379, 1.861] | **.003** | |
| Task session [8] | 1.668 [0.743, 2.592] | 1.473 [0.657, 2.290] | **< .001** | |
| Task session [9] | 1.348 [0.321, 2.378] | 1.190 [0.283, 2.101] | **.010** | |
| Task session [10] | 1.693 [0.527, 2.859] | 1.496 [0.465, 2.526] | **.004** | |

*N* = 169, 682 observations included in the model. Group [CU], Years * Group [CU], Sex [male], Study cohort [DELCODE], Distraction [no], Time of day [6AM to 12PM], and Task session [1] serve as reference points.

| | | Group-wise estimates (*b*) | Difference from MCI Aβ− |
|---|---|---|---|
| Group | CU | 0.675 [0.125, 1.226] | |
| | MCI Aβ− | 0.009 [−0.747, 0.764] | |
| | MCI Aβ+ | 0.067 [−0.720, 0.854] | 0.058 [−0.676, 0.793] |
| Years * Group | CU | −1.115 [−2.174, −0.056] | |
| | MCI Aβ− | −1.203 [−2.971, 0.565] | |
| | MCI Aβ+ | −2.401 [−4.094, −0.709] | −1.198 [−3.027, 0.631] |

*Note.* Parameters in bold are discussed in the main text. *P*-values in bold are significant at $p < .050$ uncorrected. CU = cognitively unimpaired, MCI Aβ− = mild cognitive impairment without amyloid pathology, MCI Aβ+ = mild cognitive impairment with amyloid pathology.

**Supplementary Table 7.**

Linear mixed model results for the MDT-S comparing CU and MCI groups.

| Parameter | Unstandardized estimate (b) | Standardized estimate (β) | p-value | $\eta^2_p$ |
|---|---|---|---|---|
| Intercept | 0.077 [−0.265, 0.418] | 0.248 [−0.119, 0.614] | .659 | |
| **Group [MCI]** | −0.903 [−1.385, −0.424] | −0.569 [−0.907, −0.231] | **< .001** | 0.043 |
| **Years** | −0.398 [−1.485, 0.69] | −0.061 [−0.226, 0.105] | .473 | 0.000 |
| **Years * Group [MCI]** | 0.823 [−0.244, 1.889] | 0.126 [−0.037, 0.288] | .130 | 0.004 |
| Age | −0.227 [−0.358, −0.096] | −0.186 [−0.293, −0.079] | **.001** | 0.060 |
| Sex [female] | −0.273 [−0.535, −0.011] | −0.223 [−0.437, −0.009] | **.042** | 0.023 |
| Years of education | −0.057 [−0.191, 0.076] | −0.047 [−0.156, 0.063] | .400 | 0.004 |
| Study cohort [Memory clinic] | −0.202 [−0.689, 0.285] | −0.165 [−0.563, 0.233] | .415 | 0.006 |
| Study cohort [Wisconsin] | 0.111 [−0.188, 0.409] | 0.091 [−0.154, 0.335] | .466 | |
| Concentration | 0.06 [−0.024, 0.144] | 0.049 [−0.020, 0.118] | .162 | 0.003 |
| Distraction [yes] | −0.203 [−0.464, 0.058] | −0.166 [−0.38, 0.047] | .126 | 0.003 |
| Time of day [12PM to 6PM] | 0.056 [−0.124, 0.236] | 0.046 [−0.102, 0.193] | .543 | |
| Time of day [6PM to 12AM] | −0.029 [−0.236, 0.179] | −0.023 [−0.193, 0.146] | .786 | 0.002 |
| Time of day [12AM to 6AM] | 0.164 [−0.413, 0.741] | 0.134 [−0.338, 0.605] | .576 | |
| Task session [2] | −0.359 [−0.622, −0.097] | −0.294 [−0.508, −0.079] | **.007** | |
| Task session [3] | 0.043 [−0.349, 0.434] | 0.035 [−0.285, 0.355] | .830 | |
| Task session [4] | 0.562 [0.046, 1.078] | 0.459 [0.037, 0.881] | **.033** | |
| Task session [5] | 0.581 [−0.025, 1.187] | 0.475 [−0.020, 0.970] | .060 | |
| Task session [6] | 0.282 [−0.520, 1.083] | 0.23 [−0.425, 0.886] | .490 | 0.092 |
| Task session [7] | 0.330 [−0.531, 1.191] | 0.27 [−0.434, 0.974] | .451 | |
| Task session [8] | 0.055 [−0.893, 1.003] | 0.045 [−0.73, 0.820] | .910 | |
| Task session [9] | 0.001 [−1.176, 1.179] | 0.001 [−0.962, 0.964] | .998 | |
| Task session [10] | 0.139 [−0.907, 1.186] | 0.114 [−0.741, 0.969] | .794 | |

N = 174, 717 observations included in the model. Group [CU], Years * Group [CU], Sex [male], Study cohort [DELCODE], Distraction [no], Time of day [6AM to 12PM], and Task session [1] serve as reference points. Note that the model estimating random intercepts was used as the model including both random intercepts and slopes had a singular fit.

| | | Group-wise estimates (b) |
|---|---|---|
| Group | CU | 0.000 [−0.573, 0.573] |
| | MCI | −0.903 [−1.641, −0.166] |
| Years * Group | CU | −0.398 [−1.506, 0.710] |
| | MCI | 0.425 [−1.221, 2.071] |

Note. Parameters in bold are discussed in the main text. P-values in bold are significant at p < .050 uncorrected. CU = cognitively unimpaired, MCI = mild cognitive impairment.

**Supplementary Table 8.**

Linear mixed model results for the MDT-S comparing CU, MCI Aβ− and MCI Aβ+ groups.

| Parameter | Unstandardized estimate (*b*) | Standardized estimate (*β*) | *p*-value | $\eta^2_p$ |
|---|---|---|---|---|
| Intercept | 0.078 [−0.266, 0.422] | 0.218 [−0.160, 0.596] | .655 | |
| **Group [MCI Aβ−]** | −0.999 [−1.568, −0.432] | −0.564 [−0.966, −0.163] | **.001** | 0.046 |
| **Group [MCI Aβ+]** | −0.887 [−1.469, −0.306] | −0.597 [−1.005, −0.189] | **.003** | |
| **Years** | −0.409 [−1.531, 0.713] | −0.063 [−0.235, 0.110] | .474 | 0.000 |
| **Years * Group [MCI Aβ−]** | 1.235 [−0.126, 2.595] | 0.190 [−0.019, 0.399] | .075 | 0.006 |
| **Years * Group [MCI Aβ+]** | 0.635 [−0.656, 1.924] | 0.098 [−0.101, 0.296] | .334 | |
| Age | −0.213 [−0.346, −0.079] | −0.175 [−0.285, −0.065] | **.002** | 0.051 |
| Sex [female] | −0.275 [−0.542, −0.008] | −0.226 [−0.446, −0.007] | **.044** | 0.023 |
| Years of education | −0.070 [−0.207, 0.067] | −0.058 [−0.170, 0.055] | .314 | 0.006 |
| Study cohort [Memory clinic] | −0.241 [−0.743, 0.262] | −0.198 [−0.611, 0.215] | .346 | 0.007 |
| Study cohort [Wisconsin] | 0.116 [−0.185, 0.417] | 0.096 [−0.152, 0.343] | .447 | |
| Concentration | 0.047 [−0.038, 0.133] | 0.039 [−0.031, 0.109] | .277 | 0.002 |
| Distraction [yes] | −0.182 [−0.446, 0.082] | −0.150 [−0.367, 0.067] | .175 | 0.003 |
| Time of day [12PM to 6PM] | 0.059 [−0.122, 0.240] | 0.049 [−0.101, 0.197] | .522 | |
| Time of day [6PM to 12AM] | −0.077 [−0.287, 0.133] | −0.063 [−0.236, 0.110] | .474 | 0.004 |
| Time of day [12AM to 6AM] | 0.161 [−0.410, 0.731] | 0.132 [−0.337, 0.601] | .579 | |
| Task session [2] | −0.353 [−0.621, −0.085] | −0.291 [−0.511, −0.070] | **.010** | |
| Task session [3] | 0.051 [−0.353, 0.455] | 0.042 [−0.291, 0.374] | .805 | |
| Task session [4] | 0.569 [0.034, 1.104] | 0.468 [0.028, 0.908] | **.037** | |
| Task session [5] | 0.564 [−0.065, 1.194] | 0.464 [−0.054, 0.982] | .079 | |
| Task session [6] | 0.182 [−0.659, 1.024] | 0.150 [−0.542, 0.842] | .671 | 0.090 |
| Task session [7] | 0.660 [−0.248, 1.569] | 0.543 [−0.204, 1.290] | .154 | |
| Task session [8] | 0.301 [−0.699, 1.301] | 0.247 [−0.575, 1.070] | .555 | |
| Task session [9] | 0.509 [−0.601, 1.620] | 0.419 [−0.494, 1.332] | .368 | |
| Task session [10] | 0.219 [−1.034, 1.474] | 0.180 [−0.851, 1.212] | .731 | |

*N* = 169, 682 observations included in the model. Group [CU], Years * Group [CU], Sex [male], Study cohort [DELCODE], Distraction [no], Time of day [6AM to 12PM], and Task session [1] serve as reference points. Note that the model estimating random intercepts was used as the model including both random intercepts and slopes had a singular fit.

| | | Group-wise estimates (*b*) | Difference from MCI Aβ− |
|---|---|---|---|
| | CU | 0.104 [−0.488, 0.696] | |
| Group | MCI Aβ− | −0.895 [−1.698, −0.093] | |
| | MCI Aβ+ | −0.783 [−1.618, 0.052] | 0.113 [−0.654, 0.880] |
| | CU | −0.409 [−1.555, 0.737] | |
| Years * Group | MCI Aβ− | 0.826 [−1.058, 2.709] | |
| | MCI Aβ+ | 0.225 [−1.577, 2.028] | −0.600 [−2.510, 1.310] |

*Note.* Parameters in bold are discussed in the main text. *P*-values in bold are significant at $p < .050$ uncorrected. CU = cognitively unimpaired, MCI Aβ− = mild cognitive impairment without amyloid pathology, MCI Aβ+ = mild cognitive impairment with amyloid pathology.

### Supplementary Table 9.

Linear mixed model results for the ORR comparing CU and MCI groups.

| Parameter | Unstandardized estimate (*b*) | Standardized estimate (*β*) | *p*-value | $\eta^2_p$ |
|---|---|---|---|---|
| Intercept | −0.389 [−0.718, −0.060] | −0.174 [−0.513, 0.166] | **.021** | |
| **Group [MCI]** | −1.429 [−1.911, −0.947] | −1.013 [−1.363, −0.661] | **< .001** | 0.155 |
| **Years** | −0.130 [−1.085, 0.821] | −0.020 [−0.165, 0.125] | .787 | 0.000 |
| **Years * Group [MCI]** | 0.681 [−0.261, 1.626] | 0.104 [−0.040, 0.247] | .154 | 0.007 |
| Age | −0.201 [−0.332, −0.070] | −0.162 [−0.268, −0.056] | **.003** | 0.051 |
| Sex [female] | 0.104 [−0.160, 0.368] | 0.084 [−0.129, 0.297] | .437 | 0.002 |
| Years of education | 0.002 [−0.128, 0.133] | 0.002 [−0.104, 0.107] | .977 | 0.001 |
| Study cohort [Memory clinic] | −0.114 [−0.646, 0.419] | −0.092 [−0.522, 0.339] | .673 | 0.003 |
| Study cohort [Wisconsin] | 0.098 [−0.198, 0.395] | 0.079 [−0.160, 0.319] | .514 | |
| Time to retrieval | −0.330 [−0.388, −0.273] | −0.267 [−0.313, −0.221] | **< .001** | 0.082 |
| Concentration | 0.270 [0.187, 0.353] | 0.218 [0.151, 0.285] | **< .001** | 0.053 |
| Distraction [yes] | −0.354 [−0.580, −0.128] | −0.286 [−0.468, −0.104] | **.002** | 0.013 |
| Time of day [12PM to 6PM] | 0.108 [−0.038, 0.254] | 0.087 [−0.031, 0.205] | .147 | |
| Time of day [6PM to 12AM] | 0.071 [−0.082, 0.224] | 0.057 [−0.066, 0.181] | .363 | 0.005 |
| Time of day [12AM to 6AM] | −0.086 [−0.563, 0.390] | −0.070 [−0.454, 0.315] | .723 | |
| Task session [2] | 0.253 [0.033, 0.473] | 0.204 [0.026, 0.382] | **.024** | |
| Task session [3] | 0.347 [0.012, 0.683] | 0.281 [0.010, 0.552] | **.041** | |
| Task session [4] | 0.526 [0.068, 0.985] | 0.425 [0.055, 0.796] | **.023** | |
| Task session [5] | 0.219 [−0.322, 0.762] | 0.177 [−0.260, 0.616] | .422 | |
| Task session [6] | 0.613 [−0.021, 1.247] | 0.495 [−0.017, 1.007] | .056 | 0.029 |
| Task session [7] | 0.424 [−0.323, 1.172] | 0.342 [−0.261, 0.947] | .262 | |
| Task session [8] | 0.619 [−0.212, 1.452] | 0.500 [−0.171, 1.172] | .139 | |
| Task session [9] | 0.243 [−0.692, 1.180] | 0.196 [−0.559, 0.953] | .604 | |
| Task session [10] | 0.849 [−0.179, 1.879] | 0.686 [−0.144, 1.517] | .100 | |

*N* = 165, 709 observations included in the model. Group [CU], Years * Group [CU], Sex [male], Study cohort [DELCODE], Distraction [no], Time of day [6AM to 12PM], and Task session [1] serve as reference points.

| | | Group-wise estimates (*b*) |
|---|---|---|
| Group | CU | −0.008 [−0.521, 0.504] |
| | MCI | −1.437 [−2.118, −0.757] |
| Years * Group | CU | −0.130 [−1.096, 0.836] |
| | MCI | 0.551 [−0.991, 2.093] |

*Note.* Parameters in bold are discussed in the main text. *P*-values in bold are significant at $p < .050$ uncorrected. CU = cognitively unimpaired, MCI = mild cognitive impairment.

**Supplementary Table 10.**

Linear mixed model results for the ORR comparing CU, MCI Aβ− and MCI Aβ+ groups.

| Parameter | Unstandardized estimate (*b*) | Standardized estimate (*β*) | *p*-value | $\eta^2_p$ |
|---|---|---|---|---|
| Intercept | −0.388 [−0.714, −0.063] | −0.214 [−0.552, 0.123] | **.020** | |
| **Group [MCI Aβ−]** | −1.268 [−1.812, −0.723] | −0.810 [−1.201, −0.419] | **< .001** | 0.207 |
| **Group [MCI Aβ+]** | −1.899 [−2.519, −1.283] | −1.354 [−1.797, −0.910] | **< .001** | |
| **Years** | −0.236 [−1.197, 0.723] | −0.036 [−0.181, 0.110] | .628 | 0.001 |
| **Years * Group [MCI Aβ−]** | 0.998 [−0.135, 2.126] | 0.151 [−0.020, 0.322] | .083 | 0.021 |
| **Years * Group [MCI Aβ+]** | 0.816 [−0.274, 1.921] | 0.124 [−0.042, 0.291] | .143 | |
| Age | −0.220 [−0.352, −0.089] | −0.177 [−0.282, −0.071] | **.001** | 0.059 |
| Sex [female] | 0.090 [−0.174, 0.354] | 0.072 [−0.139, 0.284] | .499 | 0.001 |
| Years of education | 0.018 [−0.113, 0.150] | 0.015 [−0.090, 0.120] | .783 | 0.000 |
| Study cohort [Memory clinic] | −0.121 [−0.669, 0.430] | −0.097 [−0.536, 0.344] | .665 | 0.003 |
| Study cohort [Wisconsin] | 0.098 [−0.194, 0.392] | 0.078 [−0.156, 0.314] | .509 | |
| Time to retrieval | −0.331 [−0.388, −0.273] | −0.265 [−0.311, −0.219] | **< .001** | 0.082 |
| Concentration | 0.263 [0.179, 0.346] | 0.210 [0.143, 0.278] | **< .001** | 0.052 |
| Distraction [yes] | −0.362 [−0.587, −0.137] | −0.290 [−0.470, −0.110] | **.002** | 0.014 |
| Time of day [12PM to 6PM] | 0.121 [−0.025, 0.268] | 0.097 [−0.020, 0.215] | .105 | |
| Time of day [6PM to 12AM] | 0.066 [−0.088, 0.221] | 0.053 [−0.071, 0.177] | .398 | 0.007 |
| Time of day [12AM to 6AM] | −0.129 [−0.604, 0.346] | −0.103 [−0.483, 0.277] | .594 | |
| Task session [2] | 0.269 [0.045, 0.493] | 0.216 [0.036, 0.395] | **.018** | |
| Task session [3] | 0.386 [0.045, 0.726] | 0.309 [0.036, 0.582] | **.026** | |
| Task session [4] | 0.594 [0.126, 1.061] | 0.476 [0.101, 0.849] | **.012** | |
| Task session [5] | 0.260 [−0.287, 0.809] | 0.208 [−0.230, 0.648] | .348 | |
| Task session [6] | 0.667 [0.022, 1.310] | 0.534 [0.018, 1.049] | **.041** | 0.031 |
| Task session [7] | 0.433 [−0.327, 1.193] | 0.347 [−0.262, 0.956] | .262 | |
| Task session [8] | 0.654 [−0.186, 1.493] | 0.523 [−0.149, 1.196] | .123 | |
| Task session [9] | 0.416 [−0.544, 1.371] | 0.333 [−0.435, 1.098] | .388 | |
| Task session [10] | 1.005 [−0.035, 2.042] | 0.805 [−0.028, 1.636] | .055 | |

*N* = 160, 675 observations included in the model. Group [CU], Years * Group [CU], Sex [male], Study cohort [DELCODE], Distraction [no], Time of day [6AM to 12PM], and Task session [1] serve as reference points.

| | | Group-wise estimates (*b*) | Difference from MCI Aβ− |
|---|---|---|---|
| Group | CU | 0.029 [−0.488, 0.545] | |
| | MCI Aβ− | −1.239 [−1.971, −0.507] | |
| | MCI Aβ+ | −1.870 [−2.658, −1.082] | −0.631 [−1.451, 0.188] |
| Years * Group | CU | −0.236 [−1.216, 0.745] | |
| | MCI Aβ− | 0.763 [−0.893, 2.419] | |
| | MCI Aβ+ | 0.581 [−1.075, 2.237] | −0.182 [−1.772, 1.409] |

*Note.* Parameters in bold are discussed in the main text. *P*-values in bold are significant at $p < .050$ uncorrected. CU = cognitively unimpaired, MCI Aβ− = mild cognitive impairment without amyloid pathology, MCI Aβ+ = mild cognitive impairment with amyloid pathology.

**Supplementary Table 11.**

Linear mixed model results for the CSR comparing CU and MCI groups.

| Parameter | Unstandardized estimate (*b*) | Standardized estimate (*β*) | *p*-value | $\eta^2_p$ |
|---|---|---|---|---|
| Intercept | −0.589 [−0.893, −0.284] | −0.358 [−0.695, −0.021] | **< .001** | |
| **Group [MCI]** | −0.688 [−1.109, −0.268] | −0.841 [−1.190, −0.492] | **.001** | 0.052 |
| **Years** | −0.121 [−0.987, 0.752] | −0.020 [−0.164, 0.125] | .784 | 0.005 |
| **Years * Group [MCI]** | −1.068 [−1.963, −0.162] | −0.178 [−0.327, −0.027] | **.020** | 0.030 |
| Age | −0.040 [−0.163, 0.082] | −0.036 [−0.143, 0.072] | .512 | 0.002 |
| Sex [female] | 0.268 [0.020, 0.515] | 0.236 [0.018, 0.453] | **.032** | 0.025 |
| Years of education | 0.123 [−0.002, 0.248] | 0.108 [−0.002, 0.218] | .055 | 0.020 |
| Study cohort [Memory clinic] | −0.003 [−0.457, 0.449] | −0.002 [−0.402, 0.394] | .990 | 0.028 |
| Study cohort [Wisconsin] | 0.324 [0.048, 0.598] | 0.285 [0.043, 0.526] | **.019** | |
| Time to retrieval | −0.277 [−0.342, −0.211] | −0.243 [−0.301, −0.186] | **< .001** | 0.092 |
| Concentration | 0.158 [0.074, 0.242] | 0.139 [0.065, 0.213] | **< .001** | 0.019 |
| Distraction [yes] | −0.233 [−0.489, 0.022] | −0.205 [−0.430, 0.020] | .072 | 0.005 |
| Time of day [12PM to 6PM] | 0.169 [0.029, 0.308] | 0.149 [0.026, 0.271] | **.017** | |
| Time of day [6PM to 12AM] | 0.089 [−0.055, 0.233] | 0.078 [−0.049, 0.205] | .223 | 0.016 |
| Time of day [12AM to 6AM] | −0.256 [−0.594, 0.08] | −0.225 [−0.522, 0.070] | .135 | |
| Task session [2] | 0.078 [−0.114, 0.271] | 0.069 [−0.101, 0.239] | .424 | |
| Task session [3] | 0.074 [−0.227, 0.373] | 0.065 [−0.199, 0.328] | .629 | |
| Task session [4] | 0.394 [−0.013, 0.799] | 0.347 [−0.011, 0.703] | .056 | |
| Task session [5] | 0.438 [−0.050, 0.922] | 0.385 [−0.044, 0.810] | .075 | |
| Task session [6] | 0.453 [−0.127, 1.027] | 0.398 [−0.111, 0.903] | .122 | 0.040 |
| Task session [7] | 0.274 [−0.411, 0.953] | 0.241 [−0.362, 0.838] | .428 | |
| Task session [8] | 0.771 [−0.002, 1.536] | 0.678 [−0.002, 1.351] | **.049** | |
| Task session [9] | 0.480 [−0.402, 1.352] | 0.422 [−0.354, 1.189] | .282 | |
| Task session [10] | 0.897 [−0.079, 1.862] | 0.789 [−0.070, 1.638] | .069 | |

*N* = 176, 753 observations included in the model. Group [CU], Years * Group [CU], Sex [male], Study cohort [DELCODE], Distraction [no], Time of day [6AM to 12PM], and Task session [1] serve as reference points.

| | | Group-wise estimates (*b*) |
|---|---|---|
| Group | CU | 0.004 [−0.480, 0.488] |
| | MCI | −0.684 [−1.303, −0.066] |
| Years * Group | CU | −0.121 [−1.007, 0.765] |
| | MCI | −1.189 [−2.591, 0.212] |

*Note.* Parameters in bold are discussed in the main text. *P*-values in bold are significant at $p < .050$ uncorrected. CU = cognitively unimpaired, MCI = mild cognitive impairment.

**Supplementary Table 12.**

Linear mixed model results for the CSR comparing CU, MCI Aβ− and MCI Aβ+ groups.

| Parameter | Unstandardized estimate (*b*) | Standardized estimate (*β*) | *p*-value | $\eta^2_p$ |
|---|---|---|---|---|
| Intercept | −0.573 [−0.874, −0.272] | −0.297 [−0.636, 0.041] | **< .001** | |
| **Group [MCI Aβ−]** | −0.823 [−1.292, −0.355] | −0.852 [−1.239, −0.466] | **.001** | 0.074 |
| **Group [MCI Aβ+]** | −0.669 [−1.183, −0.154] | −0.874 [−1.298, −0.450] | **.011** | |
| **Years** | 0.065 [−0.838, 0.975] | 0.011 [−0.138, 0.161] | .888 | 0.003 |
| **Years * Group [MCI Aβ−]** | −0.636 [−1.762, 0.511] | −0.105 [−0.290, 0.084] | .269 | 0.038 |
| **Years * Group [MCI Aβ+]** | −1.352 [−2.514, −0.179] | −0.223 [−0.414, −0.029] | **.024** | |
| Age | −0.058 [−0.179, 0.063] | −0.051 [−0.155, 0.054] | .341 | 0.005 |
| Sex [female] | 0.283 [0.039, 0.527] | 0.246 [0.034, 0.457] | **.022** | 0.029 |
| Years of education | 0.131 [0.006, 0.255] | 0.113 [0.005, 0.221] | **.040** | 0.024 |
| Study cohort [Memory clinic] | −0.160 [−0.622, 0.297] | −0.139 [−0.539, 0.258] | .492 | 0.032 |
| Study cohort [Wisconsin] | 0.306 [0.037, 0.574] | 0.265 [0.032, 0.498] | **.024** | |
| Time to retrieval | −0.262 [−0.330, −0.194] | −0.227 [−0.286, −0.169] | **< .001** | 0.080 |
| Concentration | 0.147 [0.061, 0.233] | 0.127 [0.052, 0.202] | **.001** | 0.016 |
| Distraction [yes] | −0.325 [−0.594, −0.055] | −0.281 [−0.515, −0.048] | **.017** | 0.009 |
| Time of day [12PM to 6PM] | 0.180 [0.035, 0.325] | 0.156 [0.030, 0.282] | **.015** | |
| Time of day [6PM to 12AM] | 0.100 [−0.050, 0.249] | 0.086 [−0.043, 0.216] | .189 | 0.018 |
| Time of day [12AM to 6AM] | −0.252 [−0.594, 0.089] | −0.218 [−0.515, 0.077] | .147 | |
| Task session [2] | 0.021 [−0.180, 0.222] | 0.018 [−0.156, 0.192] | .838 | |
| Task session [3] | 0.000 [−0.315, 0.314] | 0.000 [−0.273, 0.272] | .999 | |
| Task session [4] | 0.293 [−0.135, 0.719] | 0.254 [−0.117, 0.624] | .176 | |
| Task session [5] | 0.330 [−0.182, 0.838] | 0.286 [−0.158, 0.726] | .199 | |
| Task session [6] | 0.332 [−0.276, 0.933] | 0.288 [−0.239, 0.809] | .278 | 0.041 |
| Task session [7] | 0.133 [−0.592, 0.851] | 0.115 [−0.513, 0.738] | .716 | |
| Task session [8] | 0.667 [−0.154, 1.479] | 0.578 [−0.134, 1.282] | .107 | |
| Task session [9] | 0.563 [−0.387, 1.504] | 0.488 [−0.335, 1.304] | .240 | |
| Task session [10] | 1.019 [−0.033, 2.059] | 0.884 [−0.029, 1.785] | .055 | |

*N* = 171, 717 observations included in the model. Group [CU], Years * Group [CU], Sex [male], Study cohort [DELCODE], Distraction [no], Time of day [6AM to 12PM], and Task session [1] serve as reference points.

| | | Group-wise estimates (*b*) | Difference from MCI Aβ− |
|---|---|---|---|
| Group | CU | −0.115 [−0.621, 0.391] | |
| | MCI Aβ− | −0.938 [−1.614, −0.263] | |
| | MCI Aβ+ | −0.784 [−1.491, −0.077] | 0.154 [−0.507, 0.815] |
| Years * Group | CU | 0.065 [−0.861, 0.991] | |
| | MCI Aβ− | −0.571 [−2.124, 0.982] | |
| | MCI Aβ+ | −1.287 [−2.924, 0.350] | −0.716 [−2.444, 1.013] |

*Note.* Parameters in bold are discussed in the main text. *P*-values in bold are significant at $p < .050$ uncorrected. CU = cognitively unimpaired, MCI Aβ− = mild cognitive impairment without amyloid pathology, MCI Aβ+ = mild cognitive impairment with amyloid pathology.

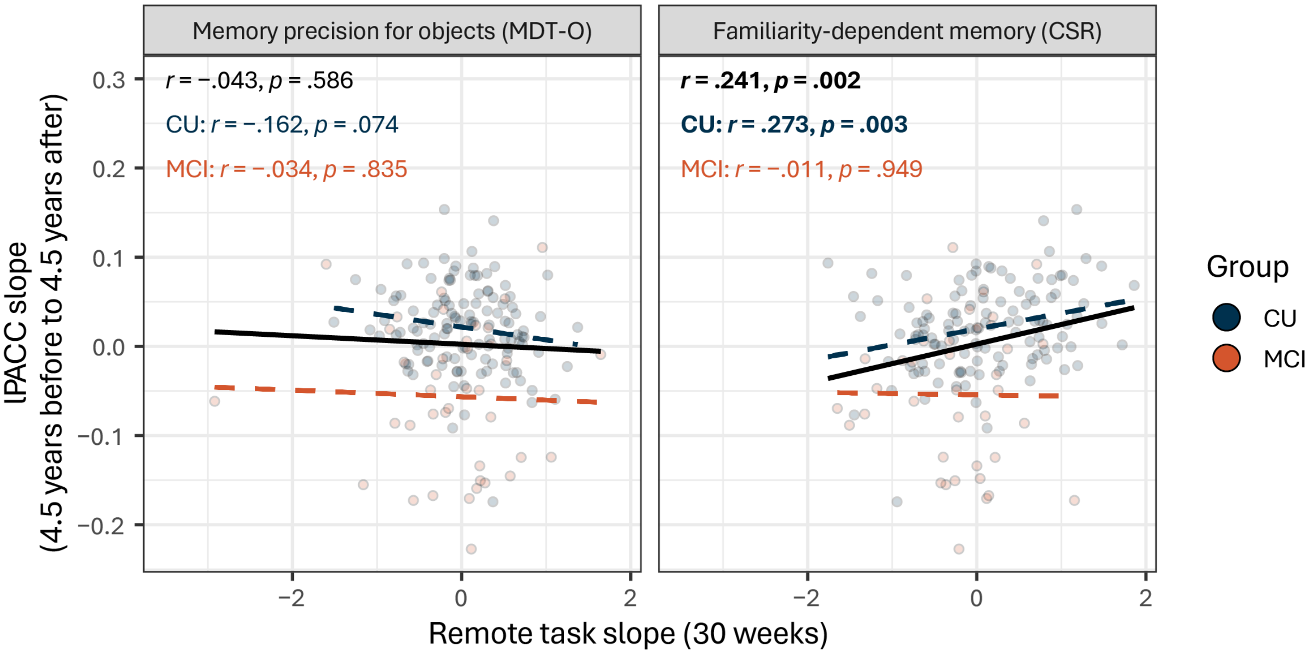


**Supplementary Fig. 6 | Correlations between remote cognitive task performance and in-person neuropsychological assessment scores.** Pearson correlations between estimated individual change on remote task performance (MDT-O and CSR only) and annual lPACC slope estimated with data from 4.5 years before and after the start of the mobile add-on study. Change in CSR showed a positive correlation with change in lPACC.

MDT-O = Mnemonic Discrimination Task for Objects; CSR = Complex Scene Recognition.

**Supplementary Table 13.**

Linear mixed model results for the lPACC comparing CU and MCI groups over one year.

| Parameter | Unstandardized estimate (*b*) | Standardized estimate (*β*) | *p*-value |
|---|---|---|---|
| Intercept | 0.133 [−0.356, 0.623] | 0.830 [0.552, 1.108] | .590 |
| **Group [MCI]** | −1.560 [−2.210, −0.905] | −0.917 [−1.288, −0.547] | **< .001** |
| **Years** | 0.008 [−0.299, 0.315] | 0.002 [−0.078, 0.082] | .957 |
| **Years * Group [MCI]** | −0.018 [−0.467, 0.443] | −0.005 [−0.121, 0.115] | .937 |
| Age | −0.160 [−0.428, 0.109] | −0.093 [−0.251, 0.064] | .240 |
| Sex [female] | 0.556 [−0.039, 1.151] | 0.326 [−0.023, 0.674] | .068 |
| Years of education | 0.407 [0.081, 0.733] | 0.239 [0.048, 0.430] | **.016** |
| Study cohort [Memory clinic] | −1.393 [−2.060, −0.727] | −0.816 [−1.207, −0.426] | **< .001** |

*N* = 46, 92 observations included in the model. Group [CU], Years * Group [CU], Sex [male], and Study cohort [Memory clinic] serve as reference points.

| | | Group-wise estimates (*b*) |
|---|---|---|
| Group | CU | −0.499 [−0.998, 0.000] |
| | MCI | −2.058 [−2.472, −1.645] |
| Years * Group | CU | 0.008 [−0.307, 0.324] |
| | MCI | −0.010 [−0.353, 0.334] |

*Note.* Parameters in bold are discussed in the main text. *P*-values in bold are significant at $p < .050$ uncorrected. CU = cognitively unimpaired, MCI = mild cognitive impairment.

### Supplementary Table 14.

Linear mixed model results for the lPACC comparing CU, MCI Aβ− and MCI Aβ+ groups over one year.

| Parameter | Unstandardized estimate (*b*) | Standardized estimate (*β*) | *p*-value |
|---|---|---|---|
| Intercept | 0.263 [−0.174, 0.699] | 0.912 [0.667, 1.157] | .235 |
| **Group [MCI Aβ−]** | −1.293 [−1.956, −0.631] | −0.698 [−1.066, −0.330] | **< .001** |
| **Group [MCI Aβ+]** | −2.172 [−2.887, −1.451] | −1.318 [−1.727, −0.911] | **< .001** |
| **Years** | 0.013 [−0.279, 0.305] | 0.003 [−0.072, 0.079] | .931 |
| **Years * Group [MCI Aβ−]** | 0.261 [−0.266, 0.789] | 0.068 [−0.069, 0.205] | .326 |
| **Years * Group [MCI Aβ+]** | −0.266 [−0.807, 0.304] | −0.069 [−0.210, 0.079] | .336 |
| Age | −0.195 [−0.432, 0.042] | −0.114 [−0.251, 0.025] | .106 |
| Sex [female] | 0.140 [−0.423, 0.703] | 0.081 [−0.246, 0.409] | .621 |
| Years of education | 0.264 [−0.030, 0.559] | 0.154 [−0.018, 0.325] | .079 |
| Study cohort [Memory clinic] | −1.199 [−1.820, −0.578] | −0.697 [−1.058, −0.336] | **< .001** |

*N* = 45, 90 observations included in the model. Group [CU], Years * Group [CU], Sex [male], and Study cohort [DELCODE] serve as reference points.

| | | Group-wise estimates (*b*) | Difference from MCI Aβ− |
|---|---|---|---|
| Group | CU | −0.425 [−0.878, 0.029] | |
| | MCI Aβ− | −1.718 [−2.284, −1.151] | |
| | MCI Aβ+ | −2.597 [−3.143, −2.051] | −0.879 [−1.863, 0.105] |
| Years * Group | CU | 0.013 [−0.290, 0.316] | |
| | MCI Aβ− | 0.274 [−0.183, 0.731] | |
| | MCI Aβ+ | −0.253 [−0.735, 0.230] | −0.527 [−1.326, 0.272] |

*Note.* Parameters in bold are discussed in the main text. *P*-values in bold are significant at $p < .050$ uncorrected. CU = cognitively unimpaired, MCI Aβ− = mild cognitive impairment without amyloid pathology, MCI Aβ+ = mild cognitive impairment with amyloid pathology.
